# Effect of Genetic and Dietary Perturbation of Glycine Metabolism on Atherosclerosis in Humans and Mice

**DOI:** 10.1101/2023.12.08.23299748

**Authors:** Subarna Biswas, James R. Hilser, Nicholas C. Woodward, Zeneng Wang, Janet Gukasyan, Ina Nemet, William S. Schwartzman, Pin Huang, Yi Han, Zachary Fouladian, Sarada Charugundla, Neal J. Spencer, Calvin Pan, W.H. Wilson Tang, Aldons J. Lusis, Stanley L. Hazen, Jaana A. Hartiala, Hooman Allayee

## Abstract

**Objective:** Epidemiological and genetic studies have reported inverse associations between circulating glycine levels and risk of coronary artery disease (CAD). However, these findings have not been consistently observed in all studies. We sought to evaluate the causal relationship between circulating glycine levels and atherosclerosis using large-scale genetic analyses in humans and dietary supplementation experiments in mice.

**Methods:** Serum glycine levels were evaluated for association with prevalent and incident CAD in the UK Biobank. A multi-ancestry genome-wide association study (GWAS) meta-analysis was carried out to identify genetic determinants for circulating glycine levels, which were then used to evaluate the causal relationship between glycine and risk of CAD by Mendelian randomization (MR). A glycine feeding study was carried out with atherosclerosis-prone apolipoprotein E deficient (*ApoE^−/−^*) mice to determine the effects of increased circulating glycine levels on amino acid metabolism, metabolic traits, and aortic lesion formation.

**Results:** Among 105,718 subjects from the UK Biobank, elevated serum glycine levels were associated with significantly reduced risk of prevalent CAD (Quintile 5 vs. Quintile 1 OR=0.76, 95% CI 0.67-0.87; P<0.0001) and incident CAD (Quintile 5 vs. Quintile 1 HR=0.70, 95% CI 0.65-0.77; P<0.0001) in models adjusted for age, sex, ethnicity, anti-hypertensive and lipid-lowering medications, blood pressure, kidney function, and diabetes. A meta-analysis of 13 GWAS datasets (total n=230,947) identified 61 loci for circulating glycine levels, of which 26 were novel. MR analyses provided modest evidence that genetically elevated glycine levels were causally associated with reduced systolic blood pressure and risk of type 2 diabetes, but did provide evidence for an association with risk of CAD. Furthermore, glycine-supplementation in *ApoE^−/−^* mice did not alter cardiometabolic traits, inflammatory biomarkers, or development of atherosclerotic lesions.

**Conclusions:** Circulating glycine levels were inversely associated with risk of prevalent and incident CAD in a large population-based cohort. While substantially expanding the genetic architecture of circulating glycine levels, MR analyses and *in vivo* feeding studies in humans and mice, respectively, did not provide evidence that the clinical association of this amino acid with CAD represents a causal relationship, despite being associated with two correlated risk factors.

## Introduction

Atherosclerotic coronary artery disease (CAD) is a complex, multi-factorial process in which many of the underlying causal biological mechanisms remain unknown^1^. For example, of the ∼300 genetic risk loci identified for CAD, only one-third harbor genes known to be involved in traditional risk factors, such as elevated lipid levels or blood pressure^2, 3^. Furthermore, lipid lowering therapies and anti-hypertensive medications are only partially effective in reducing CAD risk, and more than 50% of patients with an acute event do not exhibit these traditional risk factors^4^. Thus, there is a critical need to identify other mechanisms underlying CAD pathogenesis.

Integrating metabolomics with genetic and clinical data can offer a window into the intricate pathways governing the complex pathophysiology underlying atherosclerosis^5^. This approach has previously implicated glycine – a simple amino acid – as a potential factor in modulating risk of CAD^6, 7^. For example, dietary glycine supplementation was shown to reduce blood pressure in rats^8^ and platelet aggregation in both humans and mice^9, 10^, thus providing plausible interconnected mechanisms for the protective association observed between glycine levels and cardiovascular risk. However, recent genetic studies have provided conflicting evidence regarding the causal role of glycine in atherosclerosis^11–13^. One potential explanation for these discrepancies may be due to the pleiotropic associations that several of the genetic variants associated with glycine levels exhibit with other CAD-related traits, thus potentially violating one of the central assumptions of Mendelian randomization (MR). Consequently, it remains unclear whether the epidemiological associations observed between higher glycine levels and reduced risk of CAD represents a true inverse causal relationship. In the present study, we used complementary clinical, genetic, and dietary supplementation strategies to comprehensively evaluate the causal association of glycine with cardiovascular risk.

## Results

### Association of Serum Glycine Levels with Prevalent and Incident Risk of CAD in UK Biobank

We first leveraged data from the UK Biobank to evaluate the association of circulating glycine levels with risk of CAD. The clinical characteristics of the subjects used for these analyses for whom complete clinical, demographic, and covariate were available are shown in **Supplemental Table 1**. Among 105,718 subjects from the UK Biobank (4,099 CAD cases/101,619 controls) with metabolomics data, higher circulating glycine levels were associated with reduced risk of prevalent CAD at the time of enrollment into UK Biobank (**Table 1; Figure 1A**). This atheroprotective association was particularly evident among subjects in the highest quintiles of glycine levels compared to subjects in the first quintile (OR=0.87, 95% CI 0.78-0.96; P<0.0001 for Q4 vs. Q1 and OR=0.76, 95% CI 0.67-0.87; P<0.0001 for Q5 vs. Q1) (**Table 1; Figure 1A**). Among the 101,608 control subjects without CAD at the time of enrollment into UK Biobank and for whom complete data were available, longitudinal analyses over 5,000 days (∼13 years) of follow-up revealed that higher baseline circulating glycine levels were also associated with reduced risk of incident CAD (**Table 1; Figure 1B**). For example, subjects in the third, fourth, and fifth quintiles of glycine levels had 13-30% reduced risk of incident CAD (P<0.0001) compared to individuals with lowest levels of glycine (**Table 1; Figure 1B**).

**Figure 1.**
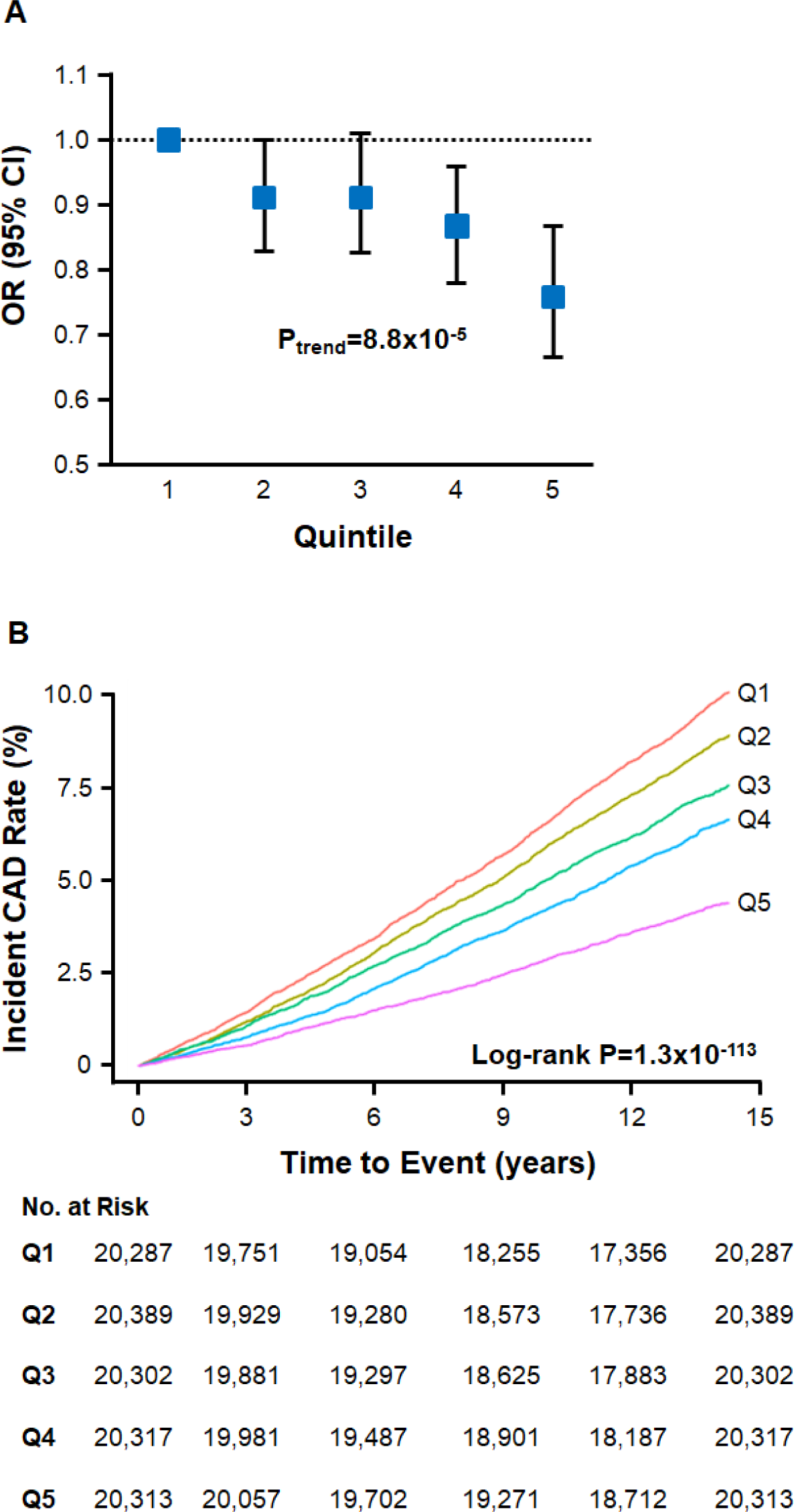
Association of Serum Glycine Levels with Risk of CAD in the UK Biobank. Individuals in the highest quintile of glycine levels had significantly reduced risk of prevalent CAD (OR=0.76, 95% CI 0.67-0.87; P<0.0001) (**A**) and incident CAD (HR=0.70, 95% CI 0.65-0.77; P<0.0001) (**B**) compared to individuals in the first quintile. P-value for trend for association with risk of prevalent CAD (**A**) and log-rank P-value for association with incident risk of CAD (**B**) across quintiles are also shown, including the number of subjects at risk of incident CAD in each quintile at baseline and the indicated follow-up time (bottom panel in **B**).

**Table 1.** Association of Serum Glycine Levels with Prevalent CAD at Time of Enrollment in UK Biobank and Incident Risk of CAD Over 5,000 Days of Follow-up.

| **Quintile for Serum Glycine Levels** | **1** | **2** | **3** | **4** | **5** |
| --- | --- | --- | --- | --- | --- |
| **Prevalent CAD** |  |  |  |  |  |
| **Range (μM)** | 0.0006-116.0 | 116.1-139.4 | 139.5-164.3 | 164.4-204.9 | 205.0-749.1 |
| **Cases/ Controls** | 1,189/19,931 | 980/20,189 | 866/20,282 | 679/20,461 | 385/20,756 |
| **OR (95% CI)** | 1 | 0.91 (0.83-1.00) | 0.91 (0.83-1.01) | **0.87 (0.78-0.96) | ***0.76 (0.67-0.87) |
| **Incident CAD** |  |  |  |  |  |
| **Range (μM)** | 0.0006-116.5 | 116.6-140.0 | 140.1-165.1 | 165.2-206.2 | 206.3-749.1 |
| **Cases/ Controls** | 1,990/18,297 | 1,783/18,606 | 1,511/18,791 | 1,322/18,995 | 895/19,418 |
| **HR (95% CI)** | 1 | 0.94 (0.88-1.01) | ***0.87 (0.81-0.92) | ***0.84 (0.78-0.90) | ***0.70 (0.65-0.77) |

Data are shown as OR or HR with 95% CIs for association of glycine quintiles with prevalent and incident CAD, respectively, where quintile 1 is the reference group. Model for prevalent CAD is adjusted for age, sex, ethnicity, anti-hypertensive medications, lipid-lowering medications, eGFR, and systolic blood pressure. Model for incident CAD is adjusted for age, sex, ethnicity, anti-hypertensive medications, lipid-lowering medications, and T2D at baseline. \*P<0.05; \*\*P<0.01; \*\*\*P<0.0001 for comparisons to quintile 1.

### Genome-Wide Meta-Analysis for Circulating Glycine Levels

We next carried out a large-scale GWAS meta-analysis to further define the genetic architecture of circulating glycine levels. In total, we combined GWAS summary statistics from 13 datasets comprising 230,947 multi-ancestry subjects (**Supplemental Table 2**). This analysis revealed 15,230 SNPs distributed among 61 loci that were associated with circulating glycine levels at the genome-wide significance threshold (P=5.0E-08) (**Figure 2A; Supplemental Table 3)**. Twenty-six of these loci were newly identified herein as being significantly associated with serum glycine levels (**Table 2; Supplemental Figure 1**), whereas the remaining 35 had been identified in previous studies^11, 12, 14^ (**Supplemental Table 3**). As expected, the larger sample size and power in our meta-analysis increased significance levels at many previously known loci, including *CPS1* (rs1047891; P=7.8E-1101) and *GLDC* (rs1801133; P=3.9E-165), which remained the two strongest genetic determinants of circulating glycine levels (**Supplemental Table 3**). In addition, several of the 26 novel loci harbored genes involved in glucose metabolism and insulin regulation, such as *IRS1* and *PPARG*, whereas genes known to be involved in the metabolism of glycine and other amino acids localized to other loci *(PCCB*, *CYP3A7*)^15, 16^ (**Supplemental Table 3; Supplemental Figure 1**). Using the absolute levels of glycine available in the UK Biobank, we also constructed a weighted genetic risk score (GRS) with all 61 SNPs, which revealed a dose-dependent increase in glycine levels as a function of carrying alleles that were associated with higher glycine (P-trend=1.57E-4). Compared to those in the bottom quintile for carrying glycine-raising alleles (range 104.2-149.0μM), serum glycine levels were significantly increased by ∼67.0±3.0μM (P=8.1E-67) among individuals in the top quintile (range 173.0-214.6μM) (**Figure 2B**).

**Figure 2.**
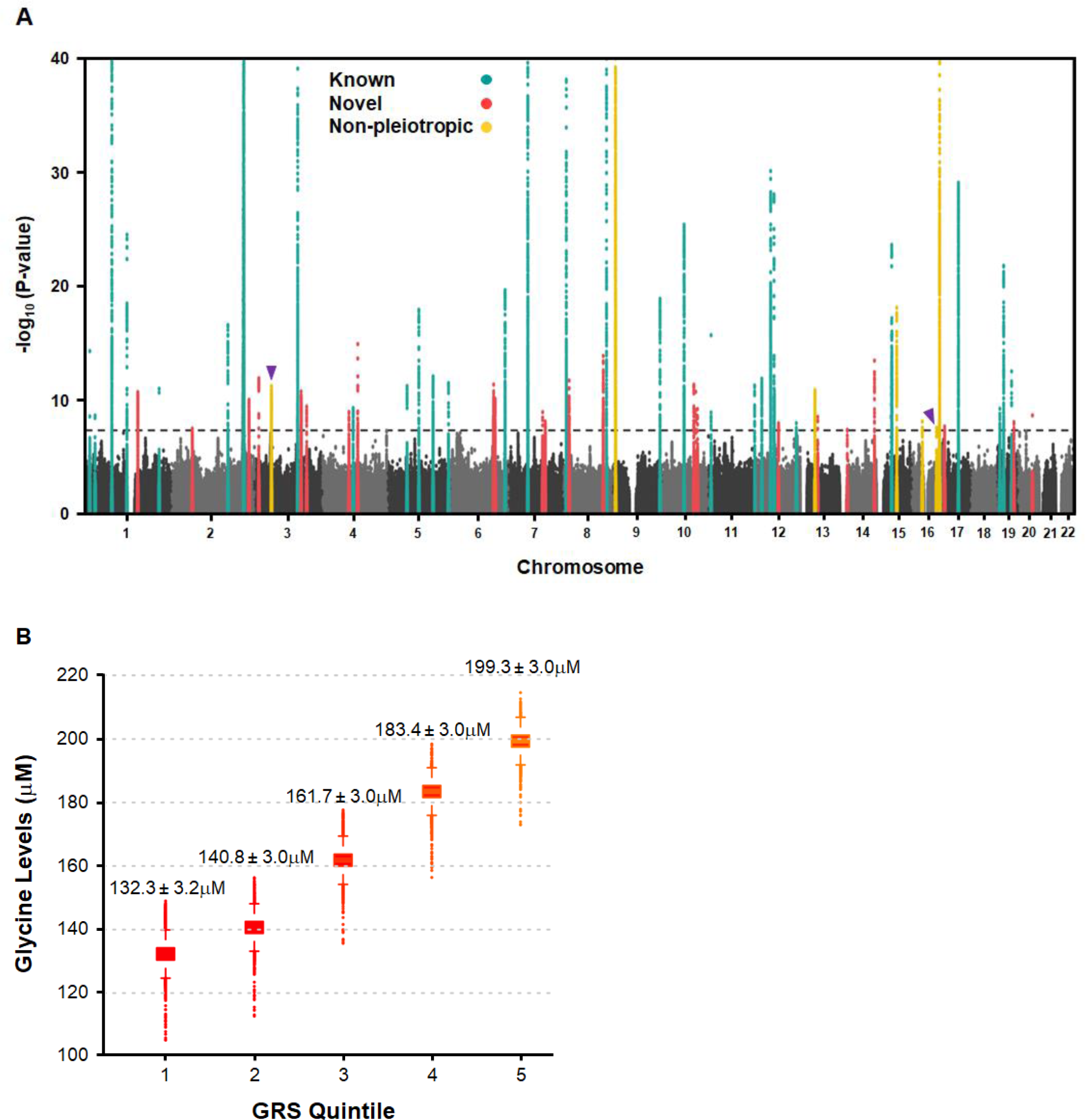
Multi-ancestry GWAS Meta-analysis and Genetic Risk Score (GRS) Analysis for Circulating Glycine Levels. (**A**) Manhattan plot shows 61 loci significantly associated with circulating glycine levels in 230,947 subjects. Novel (26) and known (35) loci are indicated by red and green dots, respectively. The seven non-pleiotropic loci are indicated by yellow dots, of which two loci on chromosomes 3 and 16 were also novel (purple arrow heads). Genome-wide thresholds for significant (P=5.0E-08) and suggestive (P=5.0E-06) association are indicated by dashed gray lines. P-values are truncated at −log_10_ (P-value)=40. (**B**) Serum glycine levels are increased as a function of quintiles of a weighted GRS constructed with the number of glycine-raising alleles carried by individuals in the UK Biobank for the 61 loci identified in the meta-analysis (n=23,283/quintile; total n=116,412). Mean glycine levels are shown for each quintile (P-trend=1.57E-4).

**Table 2.**
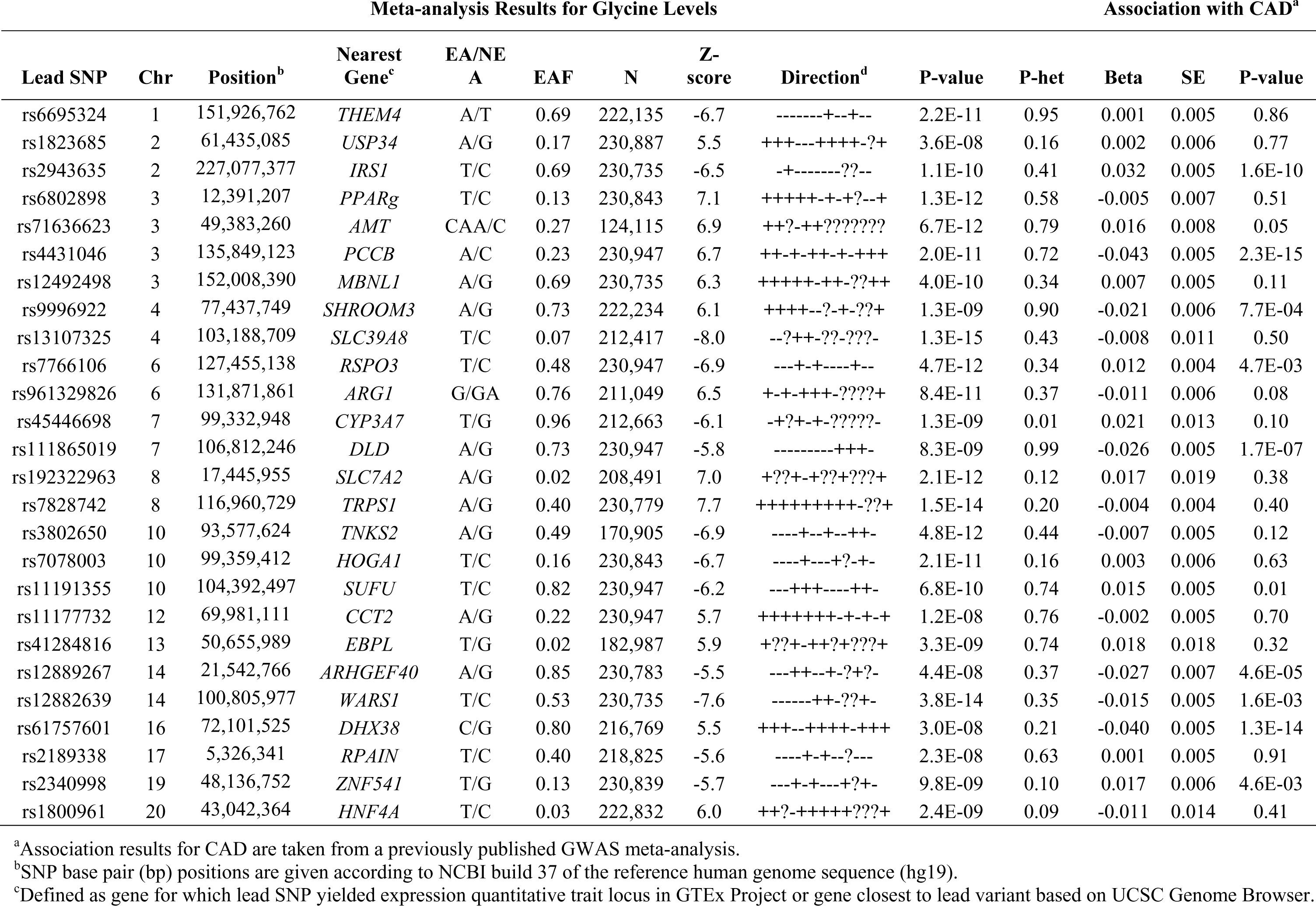
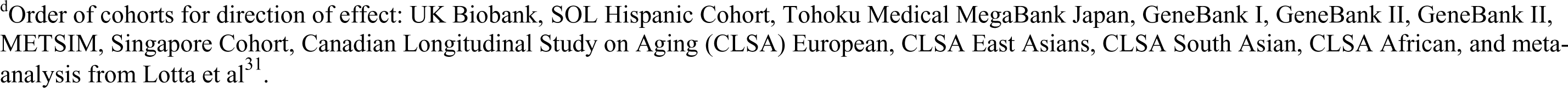
26 Novel Loci Identified for Circulating Glycine Levels.

We next carried out PheWAS analyses, which revealed that 54 of the 61 loci identified for glycine levels also exhibited pleiotropic associations with levels of other circulating metabolites or known CAD risk factors, such as lipid levels and blood pressure (**Supplemental Table 3**). Of the remaining seven loci, three harbored genes encoding components of the glycine cleavage system (*AMT, GLDC,* and *GCSH*)^17^ (**Supplemental Table 3; Supplemental Figure 1**). Genes localizing to the remaining four loci included those involved in mitochondrial regulation of oxidative stress (*VWA8*)^18^, transcription and chromosome modulators (*ZNF763, DHX38)*^19, 20^, or solute transport (*AQP9*)^21^. Altogether, the lead variants at the 61 loci explained 15.6% of the variance in glycine levels, while the seven non-pleiotropic SNPs alone explained 2.9%. Of the seven non-pleiotropic variants, one SNP (rs61757601) was also significantly associated with CAD (P=1.3E-14) (**Table 2**).

### Mendelian Randomization (MR) Analysis with Glycine-Associated SNPs and Risk of CAD

We next evaluated whether the clinical association of glycine levels with risk of CAD represented a causal relationship using various MR analyses. Variants identified from our meta-analysis for glycine levels were used as genetic instruments for the exposure and the results of a recently published large-scale GWAS for CAD^2, 3^ were used for the outcome. Weighted median and inverse variance weighted MR analyses with all 61 glycine-associated SNPs yielded modest inverse associations with CAD (OR=0.96, 95% CI 0.94-0.97; P=8.6E-07 for weighted median and OR=0.93, 95% CI 0.88-0.98; P=0.01 for inverse variance weighted) (**Figure 3A**). However, analyses with MR Egger, which takes into account pleiotropic effects of variants, did not reveal significant evidence for a causal association between genetically increased glycine levels and risk of CAD (OR=0.97, 95% CI 0.91-1.03; P=0.28) (**Figure 3A**). We also carried out weighted median and inverse variance weighted MR with the seven non-pleiotropic SNPs, which also did not yield causal evidence for the association between glycine and risk of CAD (OR=0.97, 95% CI 0.89-1.05; P=0.39 for weighted median and OR=0.93, 95% CI 0.73-1.17; P=0.52 for inverse variance weighted) (**Figure 3B**). Similarly, the most restrictive MR model that only included non-pleiotropic variants at three loci harboring genes involved in glycine cleavage system also did not reveal significant evidence for a causal association between glycine and risk of CAD (OR=0.96, 95% CI 0.89-1.05; P=0.39 for weighted median and OR=0.97, 95% CI 0.90-1.05; P=0.45 for inverse variance weighted) (**Figure 3C**). By comparison, MR analyses with the seven non-pleiotropic loci and known CAD risk factors did provide modest evidence for an inverse causal relationship between genetically increased glycine levels and systolic blood pressure and risk of T2D, but not with diastolic blood pressure, lipid levels, or T2D-related metabolic traits (**Supplemental Table 5**).

**Figure 3.**
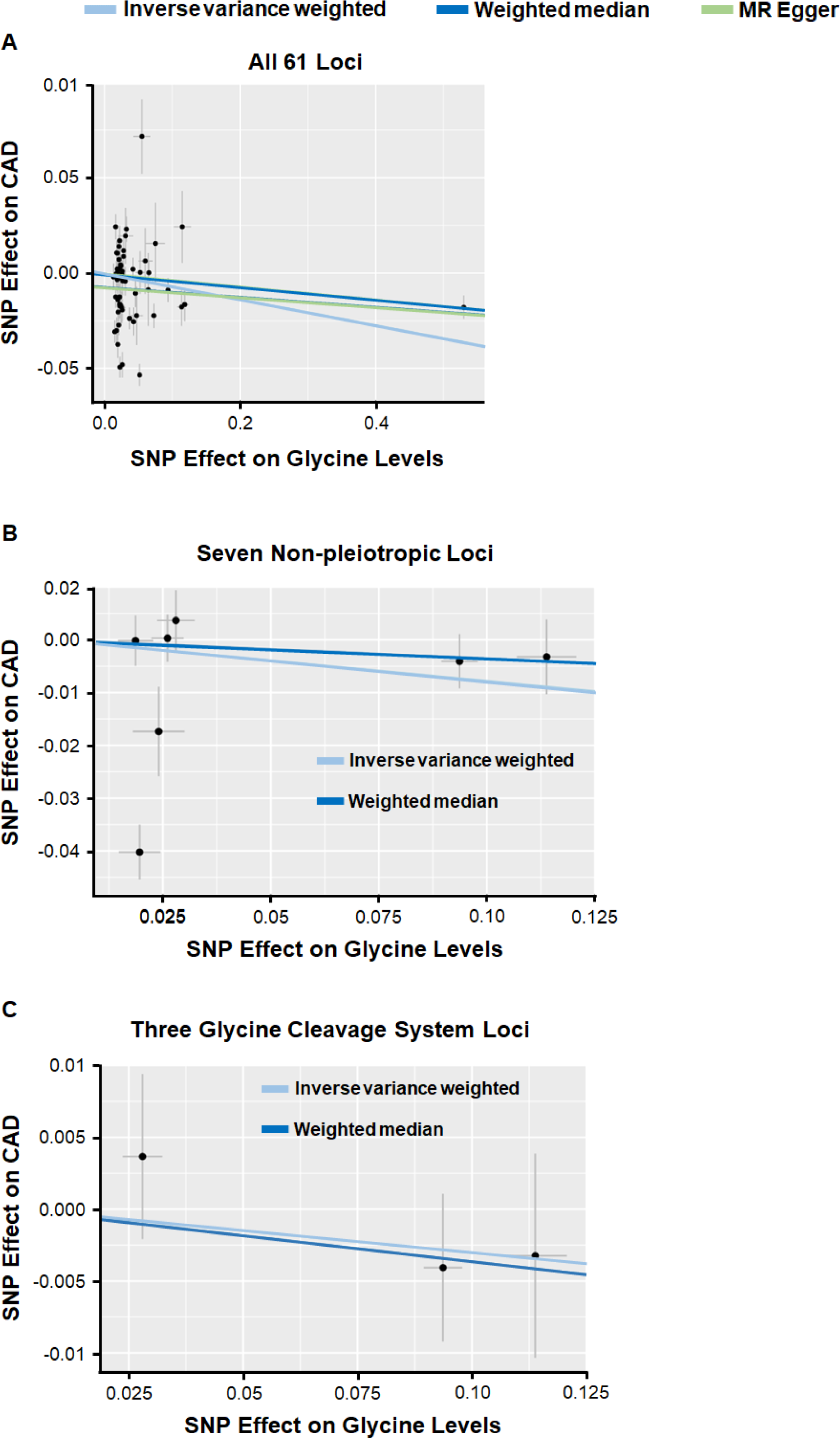
Results of MR Analyses to Evaluate Causal Association of Circulating Glycine Levels with Risk of CAD. Effect sizes of lead variants for circulating glycine levels identified in the meta-analysis (x-axis) are plotted against effect sizes for risk of CAD based on previously published summary statistics (y-axis). Slopes of the regressions are represented by the colored lines and derived from tests of MR by inverse variance weighted (light blue), weighted median (dark blue), or MR Egger (green) methods for all 61 glycine-associated loci (**A**), the seven non-pleiotropic loci (**B**), and the three non-pleiotropic loci harboring genes involved in the glycine cleavage system (**C**).

### Cardiometabolic Effects of Glycine Supplementation in Mice

To complement the human studies, we next carried out a physiologically relevant feeding study in mice to evaluate the effect of dietary glycine supplementation on cardiometabolic traits and development of atherosclerosis (**Table 3**). Amino acid-defined chow diets were developed with either 2% or 0.3% glycine content that maintained nitrogen balance (**Supplemental Table 6**), would perturb glycine levels similar to the natural variation observed in humans, and would avoid the potential confounding effects that high fat/high cholesterol atherogenic diets could have on glycine metabolism. Given these considerations, we also selected *ApoE^−/−^* mice as the mouse model with which to carry out dietary supplementation since aortic lesions develop in this strain on a chow diet. After 16 weeks of feeding, fasting glycine levels were increased in *ApoE^−/−^* mice fed the 2% glycine diet, with this elevation being highly significant in male mice and nearly significant in female mice (**Figure 4A; Table 4**). Non-fasting glycine levels were also robustly elevated in both male and female fed the glycine-enriched diet (**Figure 4B; Table 4**). Compared to the control diet, there were no significant differences in body weight, glucose and insulin-related metabolic traits, or lipid levels as a result of glycine supplementation, with the exception of a modest decrease in triglyceride levels in male mice (**Figure 4C-J; Table 3**). By comparison, male, but not female, *ApoE-/-* mice in the glycine supplementation group exhibited small differences in plasma levels of glycine-related metabolites, various amino acids, inflammatory biomarkers, and blood cell traits (**Table 4; Supplemental Tables 7-8**). However, atherosclerotic lesion area at the aortic root or along the entire aorta by *en face* analysis was not significantly different between *ApoE-/-* mice in the glycine or control diet groups (**Figure 4K-L; Table 3**).

**Figure 4.**
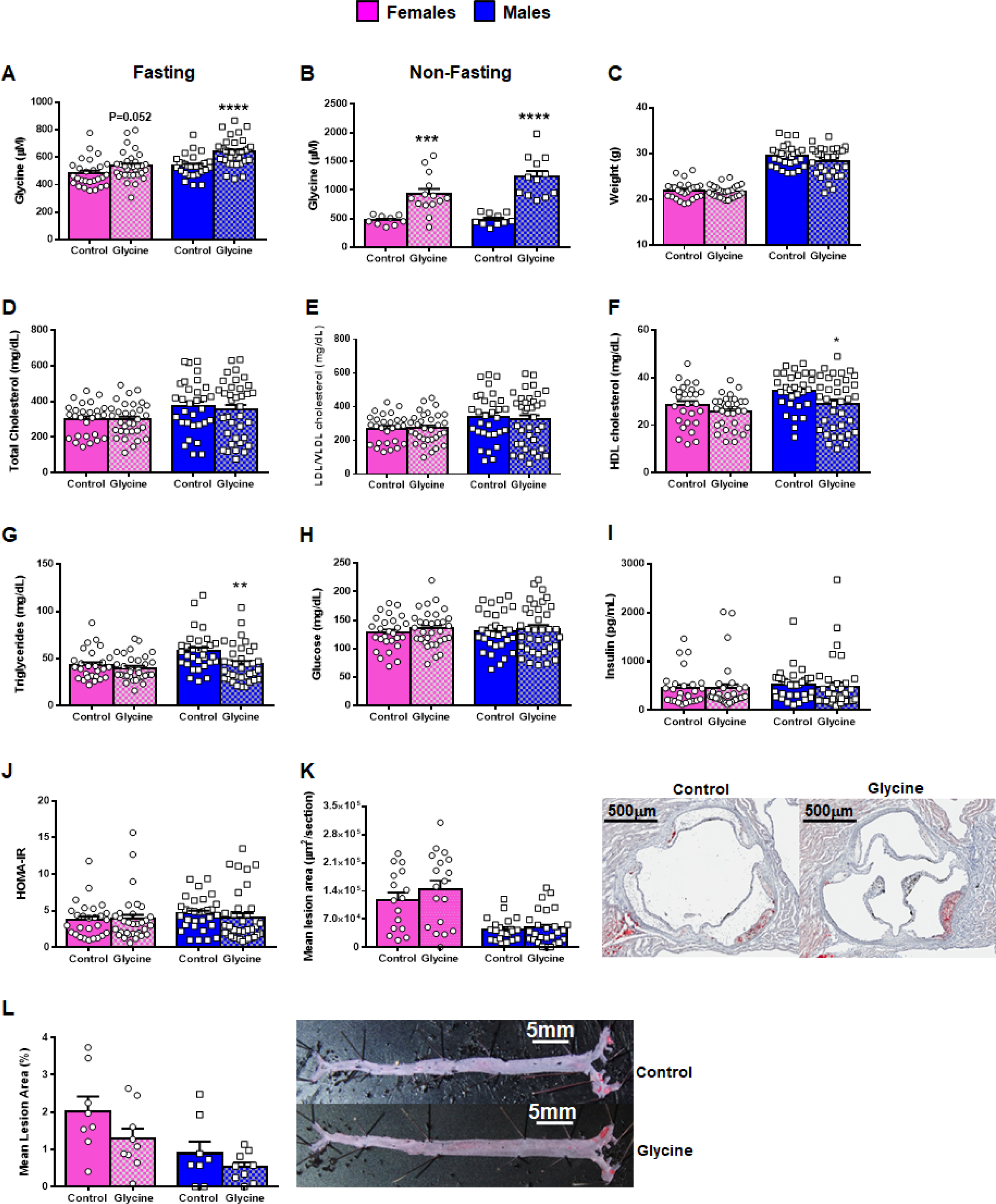
Effect of Glycine Supplementation on Plasma Cardiometabolic Traits and Atherosclerosis Development in *ApoE^−/−^* Mice. Compared to the 0.3% glycine diet (Control), mice fed the 2% glycine content diet (Glycine) had significantly higher fasting (**A**) and non-fasting (**B**) glycine levels, particularly among male mice. After 16 weeks of glycine supplementation, there were no differences in body weight (**C**) or fasting plasma levels of total cholesterol (**D**) and LDL/VLDL (**E**), whereas fasting levels of HDL cholesterol (**F**) and triglycerides (**G**) were decreased. There were also no differences with respect to metabolic traits, including fasting glucose (**H**) and insulin (**I**) levels, or HOMA-IR (**J**). Glycine supplementation did not affect atherosclerotic lesion formation assessed through serial cryosections at the aortic arch (**K**) or along the entire aorta by *en face* analysis (**L**). Representative sections of aortic lesions (**K**) and *en face* aortas stained for lipid content (**L**) are shown from female mice in the control and glycine-fed groups. Data are represented as mean ± SE. P-values are derived from t-tests carried out between control and glycine fed groups separately in males and females. *P<0.05; **P<0.01; ***P<0.001; ****P<0.0001. N=25-36 for data in panels **A** and **C-K**; n=9-14 for data in panel **B**; and n=8-9 for data in panel **L**.

**Table 3.** Effect of Dietary Glycine Supplementation on Fasting Cardiometabolic Traits and Aortic Lesion Formation in *ApoE^−/−^* Mice.

| Trait | Females |  |  | Males |  |  |
| --- | --- | --- | --- | --- | --- | --- |
|  | Control (n=27) | Glycine (n=32) | P-value | Control (n=31) | Glycine (n=36) | P-value |
| <sup>a,b</sup>Body weight (g) | 22.4 ± 0.6 | 22.2 ± 0.3 | 0.79 | 31.4 ± 0.9 | 29.7 ± 0.5 | 0.08 |
| Total cholesterol (mg/dL) | 299 ± 18 | 299 ± 17 | 0.99 | 373 ± 27 | 353 ± 27 | 0.61 |
| LDL/VLDL cholesterol (mg/dL) | 271 ± 16 | 273 ± 16 | 0.92 | 339 ± 26 | 324 ± 26 | 0.69 |
| HDL cholesterol (mg/dL) | 28 ± 2 | 26 ± 1 | 0.18 | 34 ± 1 | 29 ± 2 | 0.03 |
| Triglycerides (mg/dL) | 43 ± 3 | 40 ± 2 | 0.40 | 58 ± 4 | 44 ± 3 | 0.008 |
| <sup>c</sup>Glucose (mg/dL) | 128 ± 6 | 135 ± 6 | 0.35 | 130 ± 6 | 134 ± 7 | 0.69 |
| Insulin (pg/mL) | 453 ± 68 | 447 ± 85 | 0.96 | 523 ± 58 | 481 ± 88 | 0.70 |
| <sup>d</sup>HOMA-IR | 3.8 ± 0.5 | 3.9 ± 0.6 | 0.88 | 4.6 ± 0.4 | 4.1 ± 0.6 | 0.50 |
| <sup>e,f</sup>Aortic lesions (μm<sup>2</sup>/section) | 118,060 ± 18,477 | 144,598 ± 21,357 | 0.36 | 44,414 ± 5,967 | 48,423 ± 8,097 | 0.70 |
| <sup>g</sup>En Face lesions (%) | 2.0 ± 0.4 | 1.3 ± 0.3 | 0.14 | 0.9 ± 0.3 | 0.5 ± 0.1 | 0.26 |

<sup>a</sup>Body weight data in females on the control and glycine diets are from 25 and 30 mice, respectively.
<sup>b</sup>Body weights data in males on the control and glycine diets are from 28 and 33 mice, respectively.
<sup>c</sup>Glucose levels in male mice on the control diet are from 30 animals.
<sup>d</sup>Calculated according to the formula: [glucose mg/dL) X insulin (μIU/ml)]/405.
<sup>e</sup>Aortic lesion data in females on the control and glycine diets are from 16 and 18 mice, respectively.
<sup>f</sup>Aortic lesion data in males on the control and glycine diets are from 22 and 27 mice, respectively.
<sup>g</sup>*En face* lesion data are from 8 mice of each sex on the control diet and 9 mice of each sex on the glycine diet.

**Table 4.** Effect of Dietary Glycine Supplementation on Plasma Levels of Glycine-related Metabolites and Amino Acids in *ApoE^−/−^* Mice.

| Metabolite (μM) | Fasting |  |  |  |  |  | Non-Fasting |  |  |  |  |  |
| --- | --- | --- | --- | --- | --- | --- | --- | --- | --- | --- | --- | --- |
|  | Females |  |  | Males |  |  | Females |  |  | Males |  |  |
|  | Control (n=27) | Glycine (n=32) | P-value | Control (n=30) | Glycine (n=36) | P-value | Control (n=9) | Glycine (n=14) | P-value | Control (n=10) | Glycine (n=12) | P-value |
| Choline | 22 ± 1 | 21 ± 1 | 0.47 | 22 ± 1 | 22 ± 1 | 0.73 | 12.6 ± 1.1 | 12.2 ± 0.8 | 0.76 | 13.1 ± 1.5 | 11.3 ± 1.1 | 0.32 |
| TMAO | 2.8 ± 0.4 | 2.9 ± 0.3 | 0.87 | 1.0 ± 0.1 | 1.1 ± 0.1 | 0.69 | 5.5 ± 0.5 | 9.5 ± 1.6 | 0.06 | 2.2 ± 0.3 | 2.6 ± 0.2 | 0.35 |
| Betaine | 49 ± 3 | 50 ± 3 | 0.88 | 43 ± 3 | 45 ± 3 | 0.73 | 46 ± 3 | 47 ± 4 | 0.77 | **37 ± 5** | **23 ± 1** | **0.01** |
| Dimethylglycine | 8.8 ± 0.3 | 9.1 ± 0.4 | 0.52 | 6.4 ± 0.3 | 6.2 ± 0.2 | 0.60 | 4.1 ± 0.2 | 4.5 ± 0.4 | 0.40 | 2.5 ± 0.2 | 1.9 ± 0.1 | 0.02 |
| Glycine | 484 ± 19 | 536 ± 18 | 0.053 | **538 ± 14** | **639 ± 16** | **1.8E-05** | **472 ± 27** | **926 ± 92** | **9.6E-04** | **485 ± 33** | **1230 ± 103** | **3.5E-06** |
| <sup>a</sup>Citrulline | 91 ± 2 | 94 ± 5 | 0.57 | **77 ± 2** | **68 ± 2** | **3.4E-04** | 109 ± 5 | 102 ± 4 | 0.30 | **88 ± 3** | **75 ± 4** | **0.02** |
| Arginine | 65 ± 4 | 73 ± 3 | 0.10 | 60 ± 2 | 61 ± 3 | 0.75 | 76 ± 8 | 72 ± 5 | 0.63 | 51 ± 5 | 54 ± 5 | 0.69 |
| Creatinine | 12.1 ± 0.3 | 12.8 ± 0.3 | 0.17 | 11.4 ± 0.3 | 11.6 ± 0.4 | 0.60 | 9.5 ± 0.4 | 9.2 ± 0.3 | 0.56 | 10.6 ± 1.5 | 8.9 ± 1.4 | 0.40 |
| Ornithine | 139 ± 10 | 122 ± 6 | 0.16 | 115 ± 5 | 116 ± 7 | 0.88 | 119 ± 12 | 105 ± 10 | 0.38 | **158 ± 20** | **104 ± 10** | **0.02** |
| Lysine | 128 ± 4 | 135 ± 4 | 0.24 | 106 ± 4 | 106 ± 44 | 0.98 | 189 ± 15 | 194 ± 18 | 0.84 | **170 ± 12** | **139 ± 8** | **0.03** |
| Histidine | 131 ± 4 | 133 ± 3 | 0.78 | 126 ± 2 | 126 ± 4 | 0.88 | 139 ± 8 | 131 ± 11 | 0.60 | **156 ± 12** | **105 ± 6** | **1.0E-03** |
| Tryptophan | 119 ± 5 | 120 ± 4 | 0.87 | 79 ± 3 | 84 ± 4 | 0.32 | 156 ± 17 | 144 ± 15 | 0.63 | **114 ± 11** | **85 ± 7** | **0.03** |
| Serine | 115 ± 6 | 125 ± 7 | 0.32 | 112 ± 4 | 127 ± 6 | 0.06 | 186 ± 13 | 218 ± 21 | 0.26 | 178 ± 23 | 167 ± 11 | 0.67 |
| Proline | 150 ± 7 | 151 ± 4 | 0.91 | 129 ± 4 | 141 ± 8 | 0.21 | 241 ± 29 | 275 ± 51 | 0.63 | 293 ± 53 | 202 ± 33 | 0.14 |
| Valine | 104 ± 4 | 101 ± 3 | 0.62 | 102 ± 4 | 104 ± 4 | 0.79 | 136 ± 8 | 130 ± 11 | 0.70 | **147 ± 11** | **108 ± 6** | **4.0E-03** |
| Phenylalanine | 109 ± 3 | 109 ± 2 | 0.94 | 98 ± 2 | 104 ± 3 | 0.16 | 114 ± 12 | 98 ± 7 | 0.22 | **134 ± 18** | **75 ± 6** | **3.4E-03** |

Metabolites were measured by mass spectroscopy in plasmas obtained after either a 4 hour fast (fasting) or after two hours into the feeding cycle (non-fasting). P-values between control and glycine fed groups, separately in males and females and in fasted and non-fasting groups, were derived from 2-sided unpaired t-tests. Differences at P<0.05 are highlighted in bold. TMAO, trimethylamine *N*-oxide.

## Discussion

Multiple indirect associations have suggested that glycine levels could be a protective biomarker of CAD risk. However, genetic studies have produced equivocal evidence for a causal link between glycine and CAD, which implies that this amino acid may be correlated with other as yet unrecognized causal biomarkers, metabolites, or pathogenic mechanisms. In the present study, we observed consistent epidemiological associations between circulating glycine levels and both prevalent and incident CAD in the UK Biobank. We also conducted the largest GWAS meta-analysis of glycine levels to date by combining summary statistics from >230,000 subjects in the UK Biobank and 12 other datasets. While substantially expanding our understanding of the genetic architecture of circulating glycine levels, various types of MR analyses in humans coupled with a large and well-controlled feeding experiment did not provide evidence that glycine plays a direct causal atheroprotective role in humans or mice.

Our large-scale genetic analysis identified 61 loci significantly associated with circulating glycine levels and strengthened the association signals at 35 previously known loci. Notably, the identification of 26 novel loci further revealed the diversity of metabolic pathways associated with glycine levels. For example, two novel loci harbored genes with well-known roles in glucose metabolism (*IRS1* and *PPARG*). In addition, recent studies have implicated *HNF4A* and *PROX1-AS1* in insulin resistance and T2D^22, 23^, whereas loci harboring *FAM13A, RSPO3,* and *EBPL* have been associated with body fat distribution and hepatic steatosis^24–26^. Genes at other loci are likely involved in the synthesis or degradation of glycine itself. For instance, *HOGA1* catalyzes the breakdown of hydroxyproline into glyoxylate, which can serve as a substrate for glycine production^27^. By comparison, *DLD* mediates the oxidation and activation of the enzyme encoded by *GCSH*, which in turn provides the substrate for the *AMT* enzyme in the oxidative cleavage of glycine^28^. *GCSH*, *DLD*, and *AMT*, together with *GLDC,* are the four components of the glycine cleavage system that metabolizes glycine into ammonia and carbon dioxide^29^ for subsequent detoxification through the urea cycle. In this regard, *GLDC* was one the most strongly associated loci for glycine levels in our meta-analysis. Moreover, arginase, encoded by *ARG1*, catalyzes conversion of arginine to ornithine in the final step of the urea cycle and is one of the major routes for removal of ammonia produced through the degradation of nitrogen-containing compounds, including glycine^30^. However, the biological mechanisms underlying association of most of the loci for circulating glycine levels still remain to be determined.

A nonsynonymous Thr1405Asn substitution (rs1047891) in the gene encoding the rate-limiting enzyme of the urea cycle, *CPS1*, remained as the most significantly associated variant for glycine levels (P=7.8E-1101), consistent with prior studies^31^. The results of a recent large-scale GWAS also revealed the glycine-raising allele of rs1047891 (1405Asn) to be associated with decreased risk of CAD, although not at the genome-wide significance threshold^2^. The association of *CPS1* with risk of CAD was previously shown to be female-specific^32^ - similar to the sexually dimorphic association patterns observed with levels of various metabolites including glycine, and more recently, reduced risk of T2D, in East Asians^33^. Taken together, these observations suggest that glycine may have protective cardiometabolic properties^6, 11, 12, 30^. However, *CPS1* is one of the most pleiotropically associated loci in the genome^34^ and rs1047891 has been linked to multiple other CAD risk factors, such as trimethylamine *N*-oxide (TMAO)^32^, blood cell traits^35^, and uric acid levels^36, 37^. These observations would thus argue that the protective association of *CPS1* with risk of CAD could also be due to one or more of these other traits, either individually or collectively, rather than glycine per se. This notion is supported by the observation that the second most significantly associated locus for glycine levels (*GLDC*) was not associated with risk of CAD^2^ or T2D^38^, whereas the loci most strongly associated with CAD (*PIGV, TRIB1,* and *SLC22A3*) had smaller effect sizes on glycine levels than *CPS1* and *GLDC*. Prior MR analyses in humans with sample sizes smaller than in our present study had suggested that genetically higher glycine levels were associated with reduce risk of CAD^12, 13^. We also used various forms of MR with larger numbers of subjects and genetic instruments to evaluate whether glycine levels were causally associated with risk of CAD. When using lead variants at all 61 identified loci, MR analyses did yield evidence that genetically higher glycine levels were modestly associated with reduced risk of CAD. However, most of the glycine-associated loci (i.e., *CPS1*) had pleiotropic effects on other CAD-related traits, thus violating one of the fundamental assumptions of MR analysis. Therefore, we could not conclude, based on these data alone, that the atheroprotective association of glycine with CAD was attributable entirely to this amino acid. The lack of significant evidence for a causal association between genetically increased glycine levels and risk of CAD from the MR Egger analyses, which takes into account pleiotropic effects of variants, further suggested that glycine may not be causally associated with risk of CAD. To address this issue, we specifically carried out MR analyses with the seven glycine-associated loci for which there were no other reported associations, including the variants at three of the four loci harboring genes of the glycine cleavage complex (*AMT, GLDC,* and *GCSH*). However, MR analyses with both sets of non-pleiotropic variants also did not provide evidence for a causal relationship between higher glycine levels and lower risk of CAD. Collectively, these results do not provide compelling evidence for glycine having direct cardioprotective properties. MR analyses with the seven non-pleiotropic variants did suggest that genetically higher glycine levels may be associated with a modest causal effect on decreased systolic blood pressure and risk of T2D. These observations suggest that the inverse clinical associations we and others observe between glycine levels and risk of CAD could be indirect and possibly mediated through effects on peripheral metabolism and/or the vasculature.

To complement the human studies, we also carried out a comprehensive feeding study in *ApoE^−/−^* mice with isocaloric, amino acid-defined diets. Despite elevating both fasting and non-fasting glycine levels, particularly in males, the glycine-enriched diet did not reduce aortic lesion development. For the most part, glycine supplementation also did not change plasma levels of lipid and metabolic traits, other amino acids, inflammatory biomarkers, or blood cell parameters, particularly among female mice who are more prone to atherosclerosis than male mice. Importantly, our study was well-powered to detect differences in these phenotypes since, for example, there were at least ∼20 mice of each sex (and in some cases even >30 mice) in most of the experimental groups, including those used to evaluate aortic lesion formation. Taken together with our human studies, these data further argue against glycine having atheroprotective properties. By comparison, a previous study had suggested that glycine feeding in combination with a Western diet could mitigate atherosclerosis in *ApoE^−/−^* mice^39^. However, the effects of the diets on circulating glycine or lipid levels after supplementation were not shown and only male mice were used, with fewer animals in each experimental group^39^ than in our study. Interestingly, other rodent feeding studies have shown improvements in insulin sensitivity with glycine supplementation, although these effects were primarily observed in the context of high fat diets as well^40^. By contrast, chow diets supplemented with glycine did not lead to metabolic changes even with longer feeding durations^41^. Our results would be consistent with these latter observations since we also used chow diets with modified glycine content.

While our results point to interesting clinical and genetic associations with glycine levels, we also note certain limitations of our study. First, glycine measurements in the UK Biobank were only available at the time of enrollment and may fluctuate over time. There may also have been residual confounding in our multivariate regression models for prevalent and incident CAD, since some risk factors remain unknown or unmeasured. Furthermore, because the meta-analysis involved several different datasets, metabolomics measurements were carried out on different platforms, in either serum or plasma, and at different levels of dietary intake. However, the error stemming from these variations would likely bias the results towards the null and decrease the likelihood of identifying significant genetic associations. In addition, unlike the wide variation observed in overall glycine levels in the clinical analyses, the genetic instruments used in our MR analyses, even in aggregate and when only carried out with non-pleiotropic variants, may not have provided sufficient variation in glycine levels to detect significant causal associations with risk of CAD. By comparison, the stringent criteria we used to select genetic instruments in MR analyses resulted in evidence for causal associations between glycine and decreased systolic blood pressure and risk of T2D, consistent with prior studies^12, 14^. Last, it is also possible that our feeding study did not elevate glycine, at least with respect to fasting levels, sufficiently to reduce atherosclerosis. However, we elected to use diets that would lead to elevations of glycine levels that would be physiologically relevant to humans. In the UK Biobank, individuals in the top quintile for carrying glycine-raising alleles at all 61 identified loci had glycine levels that were on average ∼67μM higher than those in the bottom quintile. Furthermore, the 1405Asn variant of *CPS1*, which is the strongest genetic determinant of glycine levels in humans and is associated with reduced risk of CAD in women, increases glycine by ∼50μM per allele^32^. Thus, the increased glycine levels observed in our feeding study with *ApoE^−/−^* mice were comparable to the effects of naturally occurring genetic variants on glycine levels in humans.

In summary, our results provide a more complete picture of the genetic architecture of glycine metabolism in humans and offer numerous opportunities to further elucidate glycine metabolism in mammals. While not obtaining evidence for a direct causal relationship between glycine and atherosclerosis in humans or mice, our results still suggest glycine could indirectly mitigate CAD risk, possibly through metabolic processes and/or blood pressure regulation. Additional studies will be required to explore these possibilities.

## Methods

### Study Populations

The UK Biobank is a large, multi-site cohort that recruited participants between 40-69 years of age who were registered with a general practitioner of the UK National Health Service (NHS)^42^. Between 2006-2010, a total of 503,325 individuals were enrolled through 22 assessment centers in the UK. At enrollment, extensive data on demographics, ethnicity, education, lifestyle indicators, imaging of the body and brain, and disease-related outcomes were obtained through questionnaires, health records, and/or clinical evaluations. Blood samples were also collected at baseline for measurement of serum biomarkers that are either established disease risk factors or routinely measured as part of clinical evaluations. All study participants provided informed consent and the study was approved by the North West Multi-centre Research Ethics Committee. The present analyses with the UK Biobank were approved by the Institutional Review Board of USC Keck School of Medicine.

The Cleveland Clinic GeneBank study is a single site sample repository generated from ∼10,000 consecutive patients undergoing elective diagnostic coronary angiography or elective cardiac computed tomographic angiography with extensive clinical and laboratory characterization and longitudinal observation (https://clinicaltrials.gov/ct2/show/NCT00590200). Subject recruitment occurred between 2001 and 2007. Ethnicity was self-reported and information regarding demographics, medical history, and medication use was obtained by patient interviews and confirmed by chart reviews. All clinical outcome data were verified by source documentation. CAD was defined as adjudicated diagnoses of stable or unstable angina, myocardial infarction (MI) (adjudicated definition based on defined electrocardiographic changes or elevated cardiac enzymes), angiographic evidence of ≥ 50% stenosis of one or more major epicardial vessel, and/or a history of known CAD (documented MI, CAD, or history of revascularization). Plasma glycine levels at enrollment were quantified in 3,037 subjects of northern European ancestry in three batches (n=384, 882, and 1,771) by stable isotope dilution high performance liquid chromatography with online electrospray ionization tandem mass spectrometry (LC/MS)^43^. The GeneBank cohort has been used previously for discovery and replication of novel genes and risk factors for atherosclerotic disease^11, 32, 44–49^. Written informed consent was obtained from all participants prior to enrollment. The GeneBank study has been approved by the Institutional Review Board of the Cleveland Clinic and the present analyses were approved by the Institutional Review Board of USC Keck School of Medicine.

### Clinical Definitions in UK Biobank

All subjects in the UK Biobank with serum glycine levels were included in the clinical analyses with CAD regardless of ancestry. CAD cases were defined as having been assigned ICD10 codes I21, I22, I23, I25.2, I24.0, I24.8, I24.9, I25.0, I25.1, I25.4, I25.8, or I25.9, which included doctor-diagnosed and self-reported ischemic heart disease (Data-Fields 6150 and 20002, respectively) on or before the date of enrollment (Data-Field 53). Time to incident CAD was derived by calculating the number of days from the date when the participant without a prior CAD diagnosis attended the baseline assessment (Date-Field 53) to the date on which the first ICD10 code for CAD was assigned to the subject (Date-Field 41262 and 41280). Type 2 diabetes (T2D) status was defined based on ICD10 code E11 and kidney function was based on estimated glomerular filtration rate (eGFR), as calculated using serum creatinine, age, sex, and self-reported ethnic background (Data-Field 21000)^50^. Antihypertensive and lipid-lowering medication use were also based on self-reported data (Data-Fields 6153, 6177 and 20003).

### Clinical Analyses in UK Biobank

Glycine levels were derived from nuclear magnetic resonance (NMR)-based metabolomics analyses that were carried out on serum samples at the baseline blood draw from a random subset of ∼121,000 UK Biobank subjects^51^. Serum glycine levels were categorized into quintiles and evaluated for association with risk of prevalent CAD by logistic regression. This analysis included 101,619 controls and 4,099 cases with CAD at the time of enrollment in the UK Biobank and the model was adjusted for age, sex, self-reported ethnicity, anti-hypertensive medications, lipid-lowering medications, eGFR, and systolic blood pressure. Cox proportional hazards regression models were used to evaluate quintiles of glycine levels with incident risk of CAD with adjustment for age, sex, self-reported ethnicity, anti-hypertensive medications, lipid-lowering medications, and T2D at baseline. Association of serum glycine levels with risk incident CAD was evaluated only among participants who were defined as controls in the prevalent CAD logistic regression analysis and for whom complete data were available (n=101,608). Incident CAD was defined based on participants who were assigned an ICD10 code for CAD after enrollment into UK Biobank and up to October 31, 2022 (maximum follow-up period was capped at 5,000 days). Participants assigned an ICD10 code for CAD within 14 days of enrollment into UK Biobank were excluded and those who died from causes not classified under ICD10 codes for CAD were censored at the time of death. Trend and quintile comparison P-values were calculated for both logistic and time-to-event analyses. All statistical analyses were carried out with SAS 9.4 (SAS Institute, Inc, Cary, NC).

### GWAS for Circulating Glycine Levels in the Genebank and UK Biobank Cohorts

Genome-wide genotyping in GeneBank was carried out with either the Affymetrix Genome-Wide Human Array 6.0 Chip (n=3,031) or the Illumina Infinium Global Screening Array-24 v2.0 (GSA) BeadChip (n=1,728). Prior to imputation, genomic coordinates of SNPs on each genotyping platform were first converted to GRCh37/hg19. Quality control steps included removal of duplicate SNPs as well as those with call rates <97%, minor allele frequencies (MAFs) <1%, and without chromosome and base pair position. Individuals with genotype call rates <90%, of African American ancestry, and outliers from PCA analysis were also excluded, resulting in 671,968 SNPs in 2,972 participants genotyped with the Affymetrix 6.0 Chip, and 539,533 SNPs in 1,624 participants genotyped with the GSA Chip. Imputation was carried out for unmeasured SNPs on the forward (+) strand using 1000 Genomes Project (Phase 3 release, v5) and Haplotype Reference Consortium (vr1.1 2016) as reference panels through the University of Michigan Imputation Server (https://imputationserver.sph.umich.edu). After imputation, subjects with discordant sex were excluded and SNPs that had Hardy-Weinberg equilibrium P-values <0.0001, were duplicates or multiallelic, had imputation quality scores <0.3, and with MAFs <1% were removed. This resulted in 9,185,470 SNPs available for analysis in 3,037 GeneBank subjects with plasma glycine measurements. Circulating glycine levels were analyzed by linear regression with age, sex, and genotyping array as covariates using PLINK (v1.9) (http://www.cog-genomics.org/plink/1.9/)^52^.

In UK Biobank, quality control of samples, DNA variants, and imputation were performed by the Wellcome Trust Centre for Human Genetics^42^. Briefly, ∼90 million SNPs imputed from the Haplotype Reference Consortium, UK10K, and 1000 Genomes imputation were available in UK Biobank. After filtering on autosomal SNPs with INFO scores >0.8 (directly from the UK Biobank) and with MAFs >1%, 9,560,226 variants were available in 117,152 UK Biobank subjects with serum glycine measurements. A GWAS analysis was performed with BOLT-LMM (v2.3.4) using a standard (infinitesimal) mixed model to correct for population structure due to relatedness and ancestral heterogeneity, and with adjustment for age, sex, the first 20 principal components, and genotyping array^53^. The genome-wide significance thresholds for GWAS analyses in the GeneBank and UK Biobank cohorts were set at P=5.0E-08.

### Meta-analysis for Circulating Glycine Levels

A meta-analysis for circulating glycine levels was carried by combining the GWAS results generated in GeneBank and UK Biobank with publicly available summary statistics from 13 previously published datasets^11, 31, 37, 54–57^. In total, 230,947 multi-ancestry subjects from 13 datasets were included in the meta-analysis (**Supplemental Table 2**). GWAS summary statistics from Lotta et al.^31^ were first imputed with summary statistics imputation (SSimp) (v0.5.6) software^58^ using the European population from the 1000 Genomes Project (Phase 3 release, v5) as a reference panel for LD computation. After filtering on autosomal SNPs with MAF >1%, this imputation resulted in Z-scores and P-values for association of 7,667,668 SNPs with circulating glycine levels in the cohorts used by Lotta et al.^31^. We next harmonized the summary GWAS data from the remaining studies to also match the data in the GeneBank and UK Biobank cohorts and converted genomic coordinates for all datasets to GRCh37/hg19. A weighted Z-score meta-analysis was performed by combining the Z-score and P-value summary level data for SNPs that were common to at least two datasets (**Supplemental Table 1**), as implemented in METAL^59^. The genome-wide threshold for significant associations in the GWAS meta-analyses was set at P=5.0E-08. Manhattan and quantile-quantile (QQ) plots were generated using the ‘qqman’ package (v0.1.4)^60^. A locus was defined as novel if the lead SNP was in weak or no linkage disequilibrium (LD; r^2^<0.2) with genome-wide significant variants reported previously for circulating glycine levels. Functional Mapping and Annotation of Genome-Wide Association Studies (FUMA) program (v1.5.4)^61^ was used to define independent SNPs at glycine-associated loci as those that also yielded P<5.0E-08, were at least 500kb away from the lead variant, and were in low LD (r^2^<0.2) with other variants at the locus based on LD information from the 1000 Genomes Project. To determine whether loci identified for glycine levels were also associated with other clinical traits, CAD risk factors, and metabolites, a phenome-wide association study (PheWAS) was carried out using FUMA and publicly available resources, such as the GWAS Catalog (https://www.ebi.ac.uk/gwas/home), PhenoScanner^62^, and the UCSC Genome Browser (https://genome.ucsc.edu/). The significance threshold for PheWAS analyses was set at P=5.0E-08 with a LD cut off of r^2^≥0.8 for proxy SNPs.

### Genetic Risk Score Analyses

Primary level data from the UK Biobank were used to generate weighted genetic risk scores (GRS) with all 61 loci for glycine levels. For each variant, the number of alleles associated with increased glycine was multiplied by its respective effect size obtained from the GWAS analysis in the UK Biobank and summed together across all variants to generate the GRS. Association between quintiles of weighted GRS and glycine levels was tested using linear regression, with adjustment for age, sex, the first 20 principal components, and genotyping array, in R (v4.2.0)^63^.

### Mendelian Randomization (MR) Analyses

To evaluate whether glycine levels were causally associated with risk of CAD, we used the results of our GWAS analysis in the UK Biobank (since we had primary metabolomics data with absolute concentrations of glycine levels available in this large dataset) and publicly available summary results from a recently published large-scale multi-ancestry GWAS meta-analysis for CAD^2^. Effect sizes for three groups of genetic instruments for glycine levels as the exposure were taken from the GWAS results in the UK Biobank. Group 1 included variants at all 61 glycine-associated loci, Group 2 only included variants at the seven loci that did not exhibit pleiotropic associations with other traits or metabolites, and Group 3 only included variants at the three non-pleiotropic loci that harbored genes involved in the glycine cleavage system. MR analyses were carried out using weighted median, inverse variance weighted, and MR Egger methods, as implemented in the “TwoSampleMR” package^64, 65^ for R. MR analyses with the seven non-pleiotropic loci were also carried out for blood pressure, body mass index (BMI), glucose/insulin traits, circulating lipid levels, and risk of T2D using effect sizes from previously published GWAS meta-analyses^38, 66–69^, as the outcomes.

### Animal Husbandry and Glycine Supplementation

Animal studies were performed with approval and in accordance with the guidelines of the Institutional Animal Research Committees of the University of Southern California. All mice were housed in a temperature-controlled facility with a 12-h light/dark cycle and fed ad libitum with free access to water. Male and female *ApoE^−/−^* mice (stock number: 002052) were purchased from the Jackson Laboratories (Bar Harbor, Maine), bred in-house for this study, and maintained on a chow diet (PicoLab: #5053) until initiation of the glycine feeding studies. At 8-10 weeks of age, age-matched male and female *ApoE^−/−^* mice were placed on customized chow and amino acid defined diets containing either 0.3% glycine (Research Diets: A18011701, New Brunswick, NJ) or 2% glycine (Research Diets: A18011702, New Brunswick, NJ).

### Blood Measurements

Following 16 weeks of glycine supplementation, blood was collected from the submandibular vein of mice after a 12hr overnight fast and mice were euthanized. One week prior to euthanization, non-fasting blood was also obtained from a subset of mice two hours after the beginning of the feeding cycle. Plasma levels of glycine and other metabolites/amino acids were quantified by LC/MS, as described above for the GeneBank cohort. Plasma total cholesterol, high-density lipoprotein (HDL) cholesterol, triglyceride, glucose, and insulin levels were determined by enzymatic colorimetric assays, as described previously^70, 71^. Combined very low-density lipoprotein (VLDL) cholesterol and low-density lipoprotein (LDL) cholesterol levels were calculated by subtracting HDL cholesterol from total plasma cholesterol levels. Plasma insulin levels were measured in duplicate using Mouse Ultrasensitive Insulin ELISA kits (Alpco Inc: 80-INSMSU-E10, Salem, NH). Homeostasis modeling assessment insulin resistance (HOMA-IR) was calculated according to the formula: [glucose (mmol/L) X insulin (mIU/ml)]/22.5]^72^. Inflammatory cytokines were measured using a multiplexed immunoassay kit (Meso Scale Discovery, K15048G-1, Rockville, MD) and complete blood count profiles were determined using the HEMAVET^®^ 950FS Multi-species Hematology System (Drew Scientific, Miami Lakes, FL).

### Aortic Lesion and *En Face* Analyses

After euthanization, hearts were perfused through the left ventricle with approximately 15ml of phosphate-buffered saline (PBS) and characterized for atherosclerotic lesion formation, as described previously^73^. The hearts were then removed, placed in 10% formalin solution, and transferred to 30% sterile sucrose for 48hrs before being embedded in Optimal Cutting Temperature (OCT) compound (Fisher Scientific, 23-730-571). Serial interrupted 10μm thick aortic cryosections were cut starting at the origins of the aortic valve leaflets. Every 8th section was stained with Oil Red O and hematoxylin for quantification of atherosclerotic lesion area. For each mouse, total aortic lesion size was determined in blinded fashion by summing the lesion areas of 10 sections using Image-J software (NIH). *En face* analysis was carried out on a different set of euthanized mice after first perfusing the heart and aorta with 15ml of PBS, followed by a formal sucrose solution (4% paraformaldehyde/7.5% sucrose/10mM sodium phosphate buffer/2mM EDTA/20mM butylated hydroxytoluene) for 15mins, and rinsing again with 10ml of PBS. Aortas from the aortic arch to iliac bifurcation were removed carefully under a microscope (Richter Optica S6-RLT) and the surrounding adventitial fat tissue was dissected away. The aorta was then opened longitudinally from the aortic root to iliac bifurcation and pinned on a black rubber plate filled with PBS. The aortas were then incubated in 70% ethanol for 5mins, stained with Sudan-IV solution (5mg/ml Sudan-IV in 70% ethanol and 100% acetone) for 15mins, and de-stained with 80% ethanol for 3mins. Aortas were then briefly rinsed under running tap water to remove any residual ethanol, then submerged in PBS for image capture. Images were taken with a digital camera and Sudan-IV stained atherosclerotic lesion area was calculated using Image-J software. Lesion area along the aortic arch, descending aorta, and abdominal aorta was calculated as the percentage of total area. All image capture and quantitation for the *en face* analyses were done in a blinded fashion.

### Statistical Analyses

Differences in measured variables between control and glycine-supplemented mice were determined by unpaired Student’s *t*-tests (PRISM v6.01, GraphPad Software, Boston, MA). Values are expressed as mean ± SE and differences and were considered statistically significant at P<0.05 or at the significance threshold after correcting for multiple comparisons.

## Data availability

Individual level data used in the present study are available upon application to the UK Biobank (https://www.ukbiobank.ac.uk/). Summary statistics from the meta-analysis for glycine levels will be posted to a public repository. Summary statistics for glycine levels from all other datasets used in the present study are available through their respective publications. All other relevant data are available upon request from the authors.

## Data Availability

https://pheweb.org/metsim-metab/

https://jmorp.megabank.tohoku.ac.jp

https://www.ebi.ac.uk/gwas/

## Acknowledgments

This work was supported, in part, by NIH Grants R01HL133169 and R01HL148110. The GeneBank study was supported in part by NIH grants P01HL098055, P01HL076491, and R01HL103931. Mass Spectrometry instrumentation used for the GeneBank study was housed in a facility supported in part through a Shimadzu Center of Excellence award. We gratefully acknowledge the UK Biobank Resource for providing access to their data under Application Number 33307. The sponsors had no role in study design, data collection and analysis, decision to publish, or preparation of the manuscript.

## Author contributions

Concept and design: S.B., J.R.H., S.L.H., J.A.H., and H.A. Acquisition, analysis, and interpretation of data: S.B, J.R.H., N.C.W., Z.W., J.G., I.N., W.S.S., P.H., Y.H., Z.F., N.J.S., C.P., W.H.W.T., A.J.L., S.L.H., J.A.H., and H.A. Drafting of manuscript: S.B., J.R.H., and H.A. Critical revision of the manuscript for important intellectual content: All authors.

## References

1. Lusis AJ. Atherosclerosis. Nature. 2000;407:233–41.

2. Aragam KG, Jiang T, Goel A, Kanoni S, Wolford BN, Atri DS, Weeks EM, Wang M, Hindy G, Zhou W, Grace C, Roselli C, Marston NA, Kamanu FK, Surakka I, Venegas LM, Sherliker P, Koyama S, Ishigaki K, Asvold BO, Brown MR, Brumpton B, de Vries PS, Giannakopoulou O, Giardoglou P, Gudbjartsson DF, Guldener U, Haider SMI, Helgadottir A, Ibrahim M, Kastrati A, Kessler T, Kyriakou T, Konopka T, Li L, Ma L, Meitinger T, Mucha S, Munz M, Murgia F, Nielsen JB, Nothen MM, Pang S, Reinberger T, Schnitzler G, Smedley D, Thorleifsson G, von Scheidt M, Ulirsch JC, Biobank J, Epic CVD, Arnar DO, Burtt NP, Costanzo MC, Flannick J, Ito K, Jang DK, Kamatani Y, Khera AV, Komuro I, Kullo IJ, Lotta LA, Nelson CP, Roberts R, Thorgeirsson G, Thorsteinsdottir U, Webb TR, Baras A, Bjorkegren JLM, Boerwinkle E, Dedoussis G, Holm H, Hveem K, Melander O, Morrison AC, Orho-Melander M, Rallidis LS, Ruusalepp A, Sabatine MS, Stefansson K, Zalloua P, Ellinor PT, Farrall M, Danesh J, Ruff CT, Finucane HK, Hopewell JC, Clarke R, Gupta RM, Erdmann J, Samani NJ, Schunkert H, Watkins H, Willer CJ, Deloukas P, Kathiresan S, Butterworth AS and CARDIoGRAMplusC4D Consortium. Discovery and systematic characterization of risk variants and genes for coronary artery disease in over a million participants. Nat Genet. 2022;54:1803–1815.

3. Tcheandjieu C, Zhu X, Hilliard AT, Clarke SL, Napolioni V, Ma S, Lee KM, Fang H, Chen F, Lu Y, Tsao NL, Raghavan S, Koyama S, Gorman BR, Vujkovic M, Klarin D, Levin MG, Sinnott-Armstrong N, Wojcik GL, Plomondon ME, Maddox TM, Waldo SW, Bick AG, Pyarajan S, Huang J, Song R, Ho YL, Buyske S, Kooperberg C, Haessler J, Loos RJF, Do R, Verbanck M, Chaudhary K, North KE, Avery CL, Graff M, Haiman CA, Le Marchand L, Wilkens LR, Bis JC, Leonard H, Shen B, Lange LA, Giri A, Dikilitas O, Kullo IJ, Stanaway IB, Jarvik GP, Gordon AS, Hebbring S, Namjou B, Kaufman KM, Ito K, Ishigaki K, Kamatani Y, Verma SS, Ritchie MD, Kember RL, Baras A, Lotta LA, Regeneron Genetics Center, CARDIoGRAMplusC4D Consortium, Biobank Japan, Million Veteran Program, Kathiresan S, Hauser ER, Miller DR, Lee JS, Saleheen D, Reaven PD, Cho K, Gaziano JM, Natarajan P, Huffman JE, Voight BF, Rader DJ, Chang KM, Lynch JA, Damrauer SM, Wilson PWF, Tang H, Sun YV, Tsao PS, O’Donnell CJ and Assimes TL. Large-scale genome-wide association study of coronary artery disease in genetically diverse populations. Nat Med. 2022;28:1679–1692.

4. Wong ND, Zhao Y, Quek RGW, Blumenthal RS, Budoff MJ, Cushman M, Garg P, Sandfort V, Tsai M and Lopez JAG. Residual atherosclerotic cardiovascular disease risk in statin-treated adults: The Multi-Ethnic Study of Atherosclerosis. J Clin Lipidol. 2017;11:1223–1233.

5. Civelek M and Lusis AJ. Systems genetics approaches to understand complex traits. Nat Rev Genet. 2014;15:34–48.

6. Ding Y, Svingen GF, Pedersen ER, Gregory JF, Ueland PM, Tell GS and Nygard OK. Plasma glycine and risk of acute myocardial infarction in patients with suspected stable angina pectoris. J Am Heart Assoc. 2016;5.

7. Ding Y, Pedersen ER, Svingen GF, Helgeland O, Gregory JF, Loland KH, Meyer K, Tell GS, Ueland PM and Nygard OK. Methylenetetrahydrofolate dehydrogenase 1 polymorphisms modify the associations of plasma glycine and serine with risk of acute myocardial infarction in patients with stable angina pectoris in WENBIT3 (Western Norway B Vitamin Intervention Trial). Circ Cardiovasc Genet. 2016;9:541–547.

8. El Hafidi M, Perez I and Banos G. Is glycine effective against elevated blood pressure? Curr Opin Clin Nutr Metab Care. 2006;9:26–31.

9. Schemmer P, Zhong Z, Galli U, Wheeler MD, Xiangli L, Bradford BU, Conzelmann LO, Forman D, Boyer J and Thurman RG. Glycine reduces platelet aggregation. Amino Acids. 2013;44:925–31.

10. Amin AM, Sheau Chin L, Teh CH, Mostafa H, Mohamed Noor DA, Abdul Kader M, Kah Hay Y and Ibrahim B. Pharmacometabolomics analysis of plasma to phenotype clopidogrel high on treatment platelets reactivity in coronary artery disease patients. Eur J Pharm Sci. 2018;117:351–361.

11. Jia Q, Han Y, Huang P, Woodward NC, Gukasyan J, Kettunen J, Ala-Korpela M, Anufrieva O, Wang Q, Perola M, Raitakari O, Lehtimaki T, Viikari J, Jarvelin MR, Boehnke M, Laakso M, Mohlke KL, Fiehn O, Wang Z, Tang WHW, Hazen SL, Hartiala JA and Allayee H. Genetic determinants of circulating glycine levels and risk of coronary artery disease. J Am Heart Assoc. 2019;8:e011922.

12. Wittemans LBL, Lotta LA, Oliver-Williams C, Stewart ID, Surendran P, Karthikeyan S, Day FR, Koulman A, Imamura F, Zeng L, Erdmann J, Schunkert H, Khaw KT, Griffin JL, Forouhi NG, Scott RA, Wood AM, Burgess S, Howson JMM, Danesh J, Wareham NJ, Butterworth AS and Langenberg C. Assessing the causal association of glycine with risk of cardio-metabolic diseases. Nat Commun. 2019;10:1060.

13. Chang X, Wang L, Guan SP, Kennedy BK, Liu J, Khor CC, Low AF, Chan MY, Yuan JM, Koh WP, Friedlander Y, Dorajoo R and Heng CK. The association of genetically determined serum glycine with cardiovascular risk in East Asians. Nutr Metab Cardiovasc Dis. 2021;31:1840–1844.

14. Lin C, Sun Z, Mei Z, Zeng H, Zhao M, Hu J, Xia M, Huang T, Wang C, Gao X and Zheng Y. The causal associations of circulating amino acids with blood pressure: a Mendelian randomization study. BMC Med. 2022;20:414.

15. Wongkittichote P, Ah Mew N and Chapman KA. Propionyl-CoA carboxylase - A review. Mol Genet Metab. 2017;122:145–152.

16. Li H and Lampe JN. Neonatal cytochrome P450 CYP3A7: a comprehensive review of its role in development, disease, and xenobiotic metabolism. Arch Biochem Biophys. 2019;673:108078.

17. Ren J, Wang W, Nie J, Yuan W and Zeng AP. Understanding and engineering glycine cleavage system and related metabolic pathways for C1-Based biosynthesis. Adv Biochem Eng Biotechnol. 2022;180:273–298.

18. Luo M, Willis WT, Coletta DK, Langlais PR, Mengos A, Ma W, Finlayson J, Wagner GR, Shi CX and Mandarino LJ. Deletion of the mitochondrial protein VWA8 induces oxidative stress and an HNF4alpha compensatory response in hepatocytes. Biochemistry (Mosc). 2019;58:4983–4996.

19. Laan L, Klar J, Sobol M, Hoeber J, Shahsavani M, Kele M, Fatima A, Zakaria M, Anneren G, Falk A, Schuster J and Dahl N. DNA methylation changes in Down syndrome derived neural iPSCs uncover co-dysregulation of ZNF and HOX3 families of transcription factors. Clin Epigenetics. 2020;12:9.

20. Nishimura K, Cho Y, Tokunaga K, Nakao M, Tani T and Ideue T. DEAH box RNA helicase DHX38 associates with satellite I noncoding RNA involved in chromosome segregation. Genes Cells. 2019;24:585–590.

21. Shi Y, Yasui M and Hara-Chikuma M. AQP9 transports lactate in tumor-associated macrophages to stimulate an M2-like polarization that promotes colon cancer progression. Biochem Biophys Rep. 2022;31:101317.

22. Black MH, Fingerlin TE, Allayee H, Zhang W, Xiang AH, Trigo E, Hartiala J, Lehtinen AB, Haffner SM, Bergman RN, McEachin RC, Kjos SL, Lawrence JM, Buchanan TA and Watanabe RM. Evidence of interaction between PPARG2 and HNF4A contributing to variation in insulin sensitivity in Mexican Americans. Diabetes. 2008;57:1048–56.

23. Osman W, Hassoun A, Jelinek HF, Almahmeed W, Afandi B, Tay GK and Alsafar H. Genetics of type 2 diabetes and coronary artery disease and their associations with twelve cardiometabolic traits in the United Arab Emirates population. Gene. 2020;750:144722.

24. Fathzadeh M, Li J, Rao A, Cook N, Chennamsetty I, Seldin M, Zhou X, Sangwung P, Gloudemans MJ, Keller M, Attie A, Yang J, Wabitsch M, Carcamo-Orive I, Tada Y, Lusis AJ, Shin MK, Molony CM, McLaughlin T, Reaven G, Montgomery SB, Reilly D, Quertermous T, Ingelsson E and Knowles JW. FAM13A affects body fat distribution and adipocyte function. Nat Commun. 2020;11:1465.

25. Loh NY, Minchin JEN, Pinnick KE, Verma M, Todorcevic M, Denton N, Moustafa JE, Kemp JP, Gregson CL, Evans DM, Neville MJ, Small KS, McCarthy MI, Mahajan A, Rawls JF, Karpe F and Christodoulides C. RSPO3 impacts body fat distribution and regulates adipose cell biology in vitro. Nat Commun. 2020;11:2797.

26. Moebius FF, Fitzky BU, Wietzorrek G, Haidekker A, Eder A and Glossmann H. Cloning of an emopamil-binding protein (EBP)-like protein that lacks sterol delta8-delta7 isomerase activity. Biochem J. 2003;374:229–37.

27. Ahmed HA, Fadel FI, Abdel Mawla MA, Salah DM, Fathallah MG and Amr K. Next-generation sequencing in identification of pathogenic variants in primary hyperoxaluria among 21 Egyptian families: Identification of two novel AGXT gene mutations. Mol Genet Genomic Med. 2022;10:e1992.

28. Leung KY, De Castro SCP, Galea GL, Copp AJ and Greene NDE. Glycine cleavage system H protein is essential for embryonic viability, implying additional function beyond the glycine cleavage system. Front Genet. 2021;12:625120.

29. Kikuchi G, Motokawa Y, Yoshida T and Hiraga K. Glycine cleavage system: reaction mechanism, physiological significance, and hyperglycinemia. *Proceedings of the Japan Academy Series B*, Physical and biological sciences. 2008;84:246–63.

30. Zaric BL, Radovanovic JN, Gluvic Z, Stewart AJ, Essack M, Motwalli O, Gojobori T and Isenovic ER. Atherosclerosis linked to aberrant amino acid metabolism and immunosuppressive amino acid catabolizing enzymes. Front Immunol. 2020;11:551758.

31. Lotta LA, Pietzner M, Stewart ID, Wittemans LBL, Li C, Bonelli R, Raffler J, Biggs EK, Oliver-Williams C, Auyeung VPW, Luan J, Wheeler E, Paige E, Surendran P, Michelotti GA, Scott RA, Burgess S, Zuber V, Sanderson E, Koulman A, Imamura F, Forouhi NG, Khaw KT, MacTel C, Griffin JL, Wood AM, Kastenmuller G, Danesh J, Butterworth AS, Gribble FM, Reimann F, Bahlo M, Fauman E, Wareham NJ and Langenberg C. A cross-platform approach identifies genetic regulators of human metabolism and health. Nat Genet. 2021;53:54–64.

32. Hartiala JA, Tang WH, Wang Z, Crow AL, Stewart AF, Roberts R, McPherson R, Erdmann J, Willenborg C, Hazen SL and Allayee H. Genome-wide association study and targeted metabolomics identifies sex-specific association of CPS1 with coronary artery disease. Nat Commun. 2016;7:10558.

33. Spracklen CN, Horikoshi M, Kim YJ, Lin K, Bragg F, Moon S, Suzuki K, Tam CHT, Tabara Y, Kwak SH, Takeuchi F, Long J, Lim VJY, Chai JF, Chen CH, Nakatochi M, Yao J, Choi HS, Iyengar AK, Perrin HJ, Brotman SM, van de Bunt M, Gloyn AL, Below JE, Boehnke M, Bowden DW, Chambers JC, Mahajan A, McCarthy MI, Ng MCY, Petty LE, Zhang W, Morris AP, Adair LS, Akiyama M, Bian Z, Chan JCN, Chang LC, Chee ML, Chen YI, Chen YT, Chen Z, Chuang LM, Du S, Gordon-Larsen P, Gross M, Guo X, Guo Y, Han S, Howard AG, Huang W, Hung YJ, Hwang MY, Hwu CM, Ichihara S, Isono M, Jang HM, Jiang G, Jonas JB, Kamatani Y, Katsuya T, Kawaguchi T, Khor CC, Kohara K, Lee MS, Lee NR, Li L, Liu J, Luk AO, Lv J, Okada Y, Pereira MA, Sabanayagam C, Shi J, Shin DM, So WY, Takahashi A, Tomlinson B, Tsai FJ, van Dam RM, Xiang YB, Yamamoto K, Yamauchi T, Yoon K, Yu C, Yuan JM, Zhang L, Zheng W, Igase M, Cho YS, Rotter JI, Wang YX, Sheu WHH, Yokota M, Wu JY, Cheng CY, Wong TY, Shu XO, Kato N, Park KS, Tai ES, Matsuda F, Koh WP, Ma RCW, Maeda S, Millwood IY, Lee J, Kadowaki T, Walters RG, Kim BJ, Mohlke KL and Sim X. Identification of type 2 diabetes loci in 433,540 East Asian individuals. Nature. 2020;582:240–245.

34. Buniello A, MacArthur JAL, Cerezo M, Harris LW, Hayhurst J, Malangone C, McMahon A, Morales J, Mountjoy E, Sollis E, Suveges D, Vrousgou O, Whetzel PL, Amode R, Guillen JA, Riat HS, Trevanion SJ, Hall P, Junkins H, Flicek P, Burdett T, Hindorff LA, Cunningham F and Parkinson H. The NHGRI-EBI GWAS Catalog of published genome-wide association studies, targeted arrays and summary statistics 2019. Nucleic Acids Res. 2019;47:D1005–D1012.

35. Chen MH, Raffield LM, Mousas A, Sakaue S, Huffman JE, Moscati A, Trivedi B, Jiang T, Akbari P, Vuckovic D, Bao EL, Zhong X, Manansala R, Laplante V, Chen M, Lo KS, Qian H, Lareau CA, Beaudoin M, Hunt KA, Akiyama M, Bartz TM, Ben-Shlomo Y, Beswick A, Bork-Jensen J, Bottinger EP, Brody JA, van Rooij FJA, Chitrala K, Cho K, Choquet H, Correa A, Danesh J, Di Angelantonio E, Dimou N, Ding J, Elliott P, Esko T, Evans MK, Floyd JS, Broer L, Grarup N, Guo MH, Greinacher A, Haessler J, Hansen T, Howson JMM, Huang QQ, Huang W, Jorgenson E, Kacprowski T, Kahonen M, Kamatani Y, Kanai M, Karthikeyan S, Koskeridis F, Lange LA, Lehtimaki T, Lerch MM, Linneberg A, Liu Y, Lyytikainen LP, Manichaikul A, Martin HC, Matsuda K, Mohlke KL, Mononen N, Murakami Y, Nadkarni GN, Nauck M, Nikus K, Ouwehand WH, Pankratz N, Pedersen O, Preuss M, Psaty BM, Raitakari OT, Roberts DJ, Rich SS, Rodriguez BAT, Rosen JD, Rotter JI, Schubert P, Spracklen CN, Surendran P, Tang H, Tardif JC, Trembath RC, Ghanbari M, Volker U, Volzke H, Watkins NA, Zonderman AB, Program VAMV, Wilson PWF, Li Y, Butterworth AS, Gauchat JF, Chiang CWK, Li B, Loos RJF, Astle WJ, Evangelou E, van Heel DA, Sankaran VG, Okada Y, Soranzo N, Johnson AD, Reiner AP, Auer PL and Lettre G. Trans-ethnic and ancestry-specific blood-cell genetics in 746,667 individuals from 5 global populations. Cell. 2020;182:1198–1213 e14.

36. Nakatochi M, Kanai M, Nakayama A, Hishida A, Kawamura Y, Ichihara S, Akiyama M, Ikezaki H, Furusyo N, Shimizu S, Yamamoto K, Hirata M, Okada R, Kawai S, Kawaguchi M, Nishida Y, Shimanoe C, Ibusuki R, Takezaki T, Nakajima M, Takao M, Ozaki E, Matsui D, Nishiyama T, Suzuki S, Takashima N, Kita Y, Endoh K, Kuriki K, Uemura H, Arisawa K, Oze I, Matsuo K, Nakamura Y, Mikami H, Tamura T, Nakashima H, Nakamura T, Kato N, Matsuda K, Murakami Y, Matsubara T, Naito M, Kubo M, Kamatani Y, Shinomiya N, Yokota M, Wakai K, Okada Y and Matsuo H. Genome-wide meta-analysis identifies multiple novel loci associated with serum uric acid levels in Japanese individuals. Commun Biol. 2019;2:115.

37. Sakaue S, Kanai M, Tanigawa Y, Karjalainen J, Kurki M, Koshiba S, Narita A, Konuma T, Yamamoto K, Akiyama M, Ishigaki K, Suzuki A, Suzuki K, Obara W, Yamaji K, Takahashi K, Asai S, Takahashi Y, Suzuki T, Shinozaki N, Yamaguchi H, Minami S, Murayama S, Yoshimori K, Nagayama S, Obata D, Higashiyama M, Masumoto A, Koretsune Y, FinnGen, Ito K, Terao C, Yamauchi T, Komuro I, Kadowaki T, Tamiya G, Yamamoto M, Nakamura Y, Kubo M, Murakami Y, Yamamoto K, Kamatani Y, Palotie A, Rivas MA, Daly MJ, Matsuda K and Okada Y. A cross-population atlas of genetic associations for 220 human phenotypes. Nat Genet. 2021;53:1415–1424.

38. Mahajan A, Spracklen CN, Zhang W, Ng MCY, Petty LE, Kitajima H, Yu GZ, Rueger S, Speidel L, Kim YJ, Horikoshi M, Mercader JM, Taliun D, Moon S, Kwak SH, Robertson NR, Rayner NW, Loh M, Kim BJ, Chiou J, Miguel-Escalada I, Della Briotta Parolo P, Lin K, Bragg F, Preuss MH, Takeuchi F, Nano J, Guo X, Lamri A, Nakatochi M, Scott RA, Lee JJ, Huerta-Chagoya A, Graff M, Chai JF, Parra EJ, Yao J, Bielak LF, Tabara Y, Hai Y, Steinthorsdottir V, Cook JP, Kals M, Grarup N, Schmidt EM, Pan I, Sofer T, Wuttke M, Sarnowski C, Gieger C, Nousome D, Trompet S, Long J, Sun M, Tong L, Chen WM, Ahmad M, Noordam R, Lim VJY, Tam CHT, Joo YY, Chen CH, Raffield LM, Lecoeur C, Prins BP, Nicolas A, Yanek LR, Chen G, Jensen RA, Tajuddin S, Kabagambe EK, An P, Xiang AH, Choi HS, Cade BE, Tan J, Flanagan J, Abaitua F, Adair LS, Adeyemo A, Aguilar-Salinas CA, Akiyama M, Anand SS, Bertoni A, Bian Z, Bork-Jensen J, Brandslund I, Brody JA, Brummett CM, Buchanan TA, Canouil M, Chan JCN, Chang LC, Chee ML, Chen J, Chen SH, Chen YT, Chen Z, Chuang LM, Cushman M, Das SK, de Silva HJ, Dedoussis G, Dimitrov L, Doumatey AP, Du S, Duan Q, Eckardt KU, Emery LS, Evans DS, Evans MK, Fischer K, Floyd JS, Ford I, Fornage M, Franco OH, Frayling TM, Freedman BI, Fuchsberger C, Genter P, Gerstein HC, Giedraitis V, Gonzalez-Villalpando C, Gonzalez-Villalpando ME, Goodarzi MO, Gordon-Larsen P, Gorkin D, Gross M, Guo Y, Hackinger S, Han S, Hattersley AT, Herder C, Howard AG, Hsueh W, Huang M, Huang W, Hung YJ, Hwang MY, Hwu CM, Ichihara S, Ikram MA, Ingelsson M, Islam MT, Isono M, Jang HM, Jasmine F, Jiang G, Jonas JB, Jorgensen ME, Jorgensen T, Kamatani Y, Kandeel FR, Kasturiratne A, Katsuya T, Kaur V, Kawaguchi T, Keaton JM, Kho AN, Khor CC, Kibriya MG, Kim DH, Kohara K, Kriebel J, Kronenberg F, Kuusisto J, Lall K, Lange LA, Lee MS, Lee NR, Leong A, Li L, Li Y, Li-Gao R, Ligthart S, Lindgren CM, Linneberg A, Liu CT, Liu J, Locke AE, Louie T, Luan J, Luk AO, Luo X, Lv J, Lyssenko V, Mamakou V, Mani KR, Meitinger T, Metspalu A, Morris AD, Nadkarni GN, Nadler JL, Nalls MA, Nayak U, Nongmaithem SS, Ntalla I, Okada Y, Orozco L, Patel SR, Pereira MA, Peters A, Pirie FJ, Porneala B, Prasad G, Preissl S, Rasmussen-Torvik LJ, Reiner AP, Roden M, Rohde R, Roll K, Sabanayagam C, Sander M, Sandow K, Sattar N, Schonherr S, Schurmann C, Shahriar M, Shi J, Shin DM, Shriner D, Smith JA, So WY, Stancakova A, Stilp AM, Strauch K, Suzuki K, Takahashi A, Taylor KD, Thorand B, Thorleifsson G, Thorsteinsdottir U, Tomlinson B, Torres JM, Tsai FJ, Tuomilehto J, Tusie-Luna T, Udler MS, Valladares-Salgado A, van Dam RM, van Klinken JB, Varma R, Vujkovic M, Wacher-Rodarte N, Wheeler E, Whitsel EA, Wickremasinghe AR, van Dijk KW, Witte DR, Yajnik CS, Yamamoto K, Yamauchi T, Yengo L, Yoon K, Yu C, Yuan JM, Yusuf S, Zhang L, Zheng W, FinnGen, e MC, Raffel LJ, Igase M, Ipp E, Redline S, Cho YS, Lind L, Province MA, Hanis CL, Peyser PA, Ingelsson E, Zonderman AB, Psaty BM, Wang YX, Rotimi CN, Becker DM, Matsuda F, Liu Y, Zeggini E, Yokota M, Rich SS, Kooperberg C, Pankow JS, Engert JC, Chen YI, Froguel P, Wilson JG, Sheu WHH, Kardia SLR, Wu JY, Hayes MG, Ma RCW, Wong TY, Groop L, Mook-Kanamori DO, Chandak GR, Collins FS, Bharadwaj D, Pare G, Sale MM, Ahsan H, Motala AA, Shu XO, Park KS, Jukema JW, Cruz M, McKean-Cowdin R, Grallert H, Cheng CY, Bottinger EP, Dehghan A, Tai ES, Dupuis J, Kato N, Laakso M, Kottgen A, Koh WP, Palmer CNA, Liu S, Abecasis G, Kooner JS, Loos RJF, North KE, Haiman CA, Florez JC, Saleheen D, Hansen T, Pedersen O, Magi R, Langenberg C, Wareham NJ, Maeda S, Kadowaki T, Lee J, Millwood IY, Walters RG, Stefansson K, Myers SR, Ferrer J, Gaulton KJ, Meigs JB, Mohlke KL, Gloyn AL, Bowden DW, Below JE, Chambers JC, Sim X, Boehnke M, Rotter JI, McCarthy MI and Morris AP. Multi-ancestry genetic study of type 2 diabetes highlights the power of diverse populations for discovery and translation. Nat Genet. 2022;54:560–572.

39. Rom O, Liu Y, Finney AC, Ghrayeb A, Zhao Y, Shukha Y, Wang L, Rajanayake KK, Das S, Rashdan NA, Weissman N, Delgadillo L, Wen B, Garcia-Barrio MT, Aviram M, Kevil CG, Yurdagul A, Jr., Pattillo CB, Zhang J, Sun D, Hayek T, Gottlieb E, Mor I and Chen YE. Induction of glutathione biosynthesis by glycine-based treatment mitigates atherosclerosis. Redox Biol. 2022;52:102313.

40. El-Hafidi M, Franco M, Ramirez AR, Sosa JS, Flores JAP, Acosta OL, Salgado MC and Cardoso-Saldana G. Glycine increases insulin sensitivity and glutathione biosynthesis and protects against oxidative stress in a model of sucrose-induced insulin resistance. Oxid Med Cell Longev. 2018;2018:2101562.

41. Ceron E, Bernal-Alcantara D, Vanda B, Sommer B, Gonzalez-Trujano E and Alvarado-Vasquez N. Glycine supplementation during six months does not alter insulin, glucose or triglyceride plasma levels in healthy rats. Int J Vitam Nutr Res. 2021;91:451–460.

42. Bycroft C, Freeman C, Petkova D, Band G, Elliott LT, Sharp K, Motyer A, Vukcevic D, Delaneau O, O’Connell J, Cortes A, Welsh S, Young A, Effingham M, McVean G, Leslie S, Allen N, Donnelly P and Marchini J. The UK Biobank resource with deep phenotyping and genomic data. Nature. 2018;562:203–209.

43. Wang Z, Levison BS, Hazen JE, Donahue L, Li XM and Hazen SL. Measurement of trimethylamine-N-oxide by stable isotope dilution liquid chromatography tandem mass spectrometry. Anal Biochem. 2014;455:35–40.

44. Bhattacharyya T, Nicholls SJ, Topol EJ, Zhang R, Yang X, Schmitt D, Fu X, Shao M, Brennan DM, Ellis SG, Brennan ML, Allayee H, Lusis AJ and Hazen SL. Relationship of paraoxonase 1 (PON1) gene polymorphisms and functional activity with systemic oxidative stress and cardiovascular risk. JAMA. 2008;299:1265–76.

45. Hartiala J, Li D, Conti DV, Vikman S, Patel Y, Tang WH, Brennan ML, Newman JW, Stephensen CB, Armstrong P, Hazen SL and Allayee H. Genetic contribution of the leukotriene pathway to coronary artery disease. Hum Genet. 2011;129:617–27.

46. Tang WH, Hartiala J, Fan Y, Wu Y, Stewart AF, Erdmann J, Kathiresan S, Roberts R, McPherson R, Allayee H and Hazen SL. Clinical and genetic association of serum paraoxonase and arylesterase activities with cardiovascular risk. Arterioscler Thromb Vasc Biol. 2012;32:2803–2812.

47. Reiner AP, Hartiala J, Zeller T, Bis JC, Dupuis J, Fornage M, Baumert J, Kleber ME, Wild PS, Baldus S, Bielinski SJ, Fontes JD, Illig T, Keating BJ, Lange LA, Ojeda F, Muller-Nurasyid M, Munzel TF, Psaty BM, Rice K, Rotter JI, Schnabel RB, Tang WH, Thorand B, Erdmann J, Jacobs DR, Jr., Wilson JG, Koenig W, Tracy RP, Blankenberg S, Marz W, Gross MD, Benjamin EJ, Hazen SL and Allayee H. Genome-wide and gene-centric analyses of circulating myeloperoxidase levels in the CHARGE and CARe consortia. Hum Mol Genet. 2013;22:3381–93.

48. Hartiala J, Bennett BJ, Tang WH, Wang Z, Stewart AF, Roberts R, McPherson R, Lusis AJ, Hazen SL, Allayee H and Consortium C. Comparative genome-wide association studies in mice and humans for trimethylamine N-oxide, a proatherogenic metabolite of choline and L-carnitine. Arterioscler Thromb Vasc Biol. 2014;34:1307–13.

49. Hartiala JA, Han Y, Jia Q, Hilser JR, Huang P, Gukasyan J, Schwartzman WS, Cai Z, Biswas S, Tregouet DA, Smith NL, Invent Consortium, Charge Consortium Hemostasis Working Group, Genius-CHD Consortium, Seldin M, Pan C, Mehrabian M, Lusis AJ, Bazeley P, Sun YV, Liu C, Quyyumi AA, Scholz M, Thiery J, Delgado GE, Kleber ME, Marz W, Howe LJ, Asselbergs FW, van Vugt M, Vlachojannis GJ, Patel RS, Lyytikainen LP, Kahonen M, Lehtimaki T, Nieminen TVM, Kuukasjarvi P, Laurikka JO, Chang X, Heng CK, Jiang R, Kraus WE, Hauser ER, Ferguson JF, Reilly MP, Ito K, Koyama S, Kamatani Y, Komuro I, Biobank Japan, Stolze LK, Romanoski CE, Khan MD, Turner AW, Miller CL, Aherrahrou R, Civelek M, Ma L, Bjorkegren JLM, Kumar SR, Tang WHW, Hazen SL and Allayee H. Genome-wide analysis identifies novel susceptibility loci for myocardial infarction. Eur Heart J. 2021;42:919–933.

50. United States Renal Data System. USRDS 2015 Annual Data Report.Available at: http://www.usrds.org/adr.aspx.

51. Julkunen H, Cichonska A, Tiainen M, Koskela H, Nybo K, Makela V, Nokso-Koivisto J, Kristiansson K, Perola M, Salomaa V, Jousilahti P, Lundqvist A, Kangas AJ, Soininen P, Barrett JC and Wurtz P. Atlas of plasma NMR biomarkers for health and disease in 118,461 individuals from the UK Biobank. Nat Commun. 2023;14:604.

52. Chang CC, Chow CC, Tellier LC, Vattikuti S, Purcell SM and Lee JJ. Second-generation PLINK: rising to the challenge of larger and richer datasets. Gigascience. 2015;4:7.

53. Loh PR, Tucker G, Bulik-Sullivan BK, Vilhjalmsson BJ, Finucane HK, Salem RM, Chasman DI, Ridker PM, Neale BM, Berger B, Patterson N and Price AL. Efficient Bayesian mixed-model analysis increases association power in large cohorts. Nat Genet. 2015;47:284–90.

54. Kettunen J, Demirkan A, Wurtz P, Draisma HH, Haller T, Rawal R, Vaarhorst A, Kangas AJ, Lyytikainen LP, Pirinen M, Pool R, Sarin AP, Soininen P, Tukiainen T, Wang Q, Tiainen M, Tynkkynen T, Amin N, Zeller T, Beekman M, Deelen J, van Dijk KW, Esko T, Hottenga JJ, van Leeuwen EM, Lehtimaki T, Mihailov E, Rose RJ, de Craen AJ, Gieger C, Kahonen M, Perola M, Blankenberg S, Savolainen MJ, Verhoeven A, Viikari J, Willemsen G, Boomsma DI, van Duijn CM, Eriksson J, Jula A, Jarvelin MR, Kaprio J, Metspalu A, Raitakari O, Salomaa V, Slagboom PE, Waldenberger M, Ripatti S and Ala-Korpela M. Genome-wide study for circulating metabolites identifies 62 loci and reveals novel systemic effects of LPA. Nat Commun. 2016;7:11122.

55. Chai JF, Raichur S, Khor IW, Torta F, Chew WS, Herr DR, Ching J, Kovalik JP, Khoo CM, Wenk MR, Tai ES and Sim X. Associations with metabolites in Chinese suggest new metabolic roles in Alzheimer’s and Parkinson’s diseases. Hum Mol Genet. 2020;29:189–201.

56. Feofanova EV, Chen H, Dai Y, Jia P, Grove ML, Morrison AC, Qi Q, Daviglus M, Cai J, North KE, Laurie CC, Kaplan RC, Boerwinkle E and Yu B. A genome-wide association study discovers 46 loci of the human metabolome in the Hispanic community health study/study of Latinos. Am J Hum Genet. 2020;107:849–863.

57. Chen Y, Lu T, Pettersson-Kymmer U, Stewart ID, Butler-Laporte G, Nakanishi T, Cerani A, Liang KYH, Yoshiji S, Willett JDS, Su CY, Raina P, Greenwood CMT, Farjoun Y, Forgetta V, Langenberg C, Zhou S, Ohlsson C and Richards JB. Genomic atlas of the plasma metabolome prioritizes metabolites implicated in human diseases. Nat Genet. 2023;55:44–53.

58. Rueger S, McDaid A and Kutalik Z. Evaluation and application of summary statistic imputation to discover new height-associated loci. PLoS Genet. 2018;14:e1007371.

59. Willer CJ, Li Y and Abecasis GR. METAL: fast and efficient meta-analysis of genomewide association scans. Bioinformatics. 2010;26:2190–1.

60. Turner SD. qqman: an R package for visualizing GWAS results using Q-Q and manhattan plots. Journal of Open Source Software. 2018;3:731.

61. Watanabe K, Taskesen E, van Bochoven A and Posthuma D. Functional mapping and annotation of genetic associations with FUMA. Nat Commun. 2017;8:1826.

62. Kamat MA, Blackshaw JA, Young R, Surendran P, Burgess S, Danesh J, Butterworth AS and Staley JR. PhenoScanner V2: an expanded tool for searching human genotype-phenotype associations. Bioinformatics. 2019;35:4851–4853.

63. R: A language and environment for statistical computing. [computer program]. Vienna, Austria: R Foundation for Statistical Computing; 2022.

64. Hemani G, Tilling K and Davey Smith G. Orienting the causal relationship between imprecisely measured traits using GWAS summary data. PLoS Genet. 2017;13:e1007081.

65. Hemani G, Zheng J, Elsworth B, Wade KH, Haberland V, Baird D, Laurin C, Burgess S, Bowden J, Langdon R, Tan VY, Yarmolinsky J, Shihab HA, Timpson NJ, Evans DM, Relton C, Martin RM, Davey Smith G, Gaunt TR and Haycock PC. The MR-Base platform supports systematic causal inference across the human phenome. Elife. 2018;7:e34408.

66. Evangelou E, Warren HR, Mosen-Ansorena D, Mifsud B, Pazoki R, Gao H, Ntritsos G, Dimou N, Cabrera CP, Karaman I, Ng FL, Evangelou M, Witkowska K, Tzanis E, Hellwege JN, Giri A, Velez Edwards DR, Sun YV, Cho K, Gaziano JM, Wilson PWF, Tsao PS, Kovesdy CP, Esko T, Magi R, Milani L, Almgren P, Boutin T, Debette S, Ding J, Giulianini F, Holliday EG, Jackson AU, Li-Gao R, Lin WY, Luan J, Mangino M, Oldmeadow C, Prins BP, Qian Y, Sargurupremraj M, Shah N, Surendran P, Theriault S, Verweij N, Willems SM, Zhao JH, Amouyel P, Connell J, de Mutsert R, Doney ASF, Farrall M, Menni C, Morris AD, Noordam R, Pare G, Poulter NR, Shields DC, Stanton A, Thom S, Abecasis G, Amin N, Arking DE, Ayers KL, Barbieri CM, Batini C, Bis JC, Blake T, Bochud M, Boehnke M, Boerwinkle E, Boomsma DI, Bottinger EP, Braund PS, Brumat M, Campbell A, Campbell H, Chakravarti A, Chambers JC, Chauhan G, Ciullo M, Cocca M, Collins F, Cordell HJ, Davies G, de Borst MH, de Geus EJ, Deary IJ, Deelen J, Del Greco MF, Demirkale CY, Dorr M, Ehret GB, Elosua R, Enroth S, Erzurumluoglu AM, Ferreira T, Franberg M, Franco OH, Gandin I, Gasparini P, Giedraitis V, Gieger C, Girotto G, Goel A, Gow AJ, Gudnason V, Guo X, Gyllensten U, Hamsten A, Harris TB, Harris SE, Hartman CA, Havulinna AS, Hicks AA, Hofer E, Hofman A, Hottenga JJ, Huffman JE, Hwang SJ, Ingelsson E, James A, Jansen R, Jarvelin MR, Joehanes R, Johansson A, Johnson AD, Joshi PK, Jousilahti P, Jukema JW, Jula A, Kahonen M, Kathiresan S, Keavney BD, Khaw KT, Knekt P, Knight J, Kolcic I, Kooner JS, Koskinen S, Kristiansson K, Kutalik Z, Laan M, Larson M, Launer LJ, Lehne B, Lehtimaki T, Liewald DCM, Lin L, Lind L, Lindgren CM, Liu Y, Loos RJF, Lopez LM, Lu Y, Lyytikainen LP, Mahajan A, Mamasoula C, Marrugat J, Marten J, Milaneschi Y, Morgan A, Morris AP, Morrison AC, Munson PJ, Nalls MA, Nandakumar P, Nelson CP, Niiranen T, Nolte IM, Nutile T, Oldehinkel AJ, Oostra BA, O’Reilly PF, Org E, Padmanabhan S, Palmas W, Palotie A, Pattie A, Penninx B, Perola M, Peters A, Polasek O, Pramstaller PP, Nguyen QT, Raitakari OT, Ren M, Rettig R, Rice K, Ridker PM, Ried JS, Riese H, Ripatti S, Robino A, Rose LM, Rotter JI, Rudan I, Ruggiero D, Saba Y, Sala CF, Salomaa V, Samani NJ, Sarin AP, Schmidt R, Schmidt H, Shrine N, Siscovick D, Smith AV, Snieder H, Sober S, Sorice R, Starr JM, Stott DJ, Strachan DP, Strawbridge RJ, Sundstrom J, Swertz MA, Taylor KD, Teumer A, Tobin MD, Tomaszewski M, Toniolo D, Traglia M, Trompet S, Tuomilehto J, Tzourio C, Uitterlinden AG, Vaez A, van der Most PJ, van Duijn CM, Vergnaud AC, Verwoert GC, Vitart V, Volker U, Vollenweider P, Vuckovic D, Watkins H, Wild SH, Willemsen G, Wilson JF, Wright AF, Yao J, Zemunik T, Zhang W, Attia JR, Butterworth AS, Chasman DI, Conen D, Cucca F, Danesh J, Hayward C, Howson JMM, Laakso M, Lakatta EG, Langenberg C, Melander O, Mook-Kanamori DO, Palmer CNA, Risch L, Scott RA, Scott RJ, Sever P, Spector TD, van der Harst P, Wareham NJ, Zeggini E, Levy D, Munroe PB, Newton-Cheh C, Brown MJ, Metspalu A, Hung AM, O’Donnell CJ, Edwards TL, Psaty BM, Tzoulaki I, Barnes MR, Wain LV, Elliott P, Caulfield MJ and Million Veteran Program. Genetic analysis of over 1 million people identifies 535 new loci associated with blood pressure traits. Nat Genet. 2018;50:1412–1425.

67. Yengo L, Sidorenko J, Kemper KE, Zheng Z, Wood AR, Weedon MN, Frayling TM, Hirschhorn J, Yang J, Visscher PM and Consortium G. Meta-analysis of genome-wide association studies for height and body mass index in approximately 700000 individuals of European ancestry. Hum Mol Genet. 2018;27:3641–3649.

68. Chen J, Spracklen CN, Marenne G, Varshney A, Corbin LJ, Luan J, Willems SM, Wu Y, Zhang X, Horikoshi M, Boutin TS, Magi R, Waage J, Li-Gao R, Chan KHK, Yao J, Anasanti MD, Chu AY, Claringbould A, Heikkinen J, Hong J, Hottenga JJ, Huo S, Kaakinen MA, Louie T, Marz W, Moreno-Macias H, Ndungu A, Nelson SC, Nolte IM, North KE, Raulerson CK, Ray D, Rohde R, Rybin D, Schurmann C, Sim X, Southam L, Stewart ID, Wang CA, Wang Y, Wu P, Zhang W, Ahluwalia TS, Appel EVR, Bielak LF, Brody JA, Burtt NP, Cabrera CP, Cade BE, Chai JF, Chai X, Chang LC, Chen CH, Chen BH, Chitrala KN, Chiu YF, de Haan HG, Delgado GE, Demirkan A, Duan Q, Engmann J, Fatumo SA, Gayan J, Giulianini F, Gong JH, Gustafsson S, Hai Y, Hartwig FP, He J, Heianza Y, Huang T, Huerta-Chagoya A, Hwang MY, Jensen RA, Kawaguchi T, Kentistou KA, Kim YJ, Kleber ME, Kooner IK, Lai S, Lange LA, Langefeld CD, Lauzon M, Li M, Ligthart S, Liu J, Loh M, Long J, Lyssenko V, Mangino M, Marzi C, Montasser ME, Nag A, Nakatochi M, Noce D, Noordam R, Pistis G, Preuss M, Raffield L, Rasmussen-Torvik LJ, Rich SS, Robertson NR, Rueedi R, Ryan K, Sanna S, Saxena R, Schraut KE, Sennblad B, Setoh K, Smith AV, Sparso T, Strawbridge RJ, Takeuchi F, Tan J, Trompet S, van den Akker E, van der Most PJ, Verweij N, Vogel M, Wang H, Wang C, Wang N, Warren HR, Wen W, Wilsgaard T, Wong A, Wood AR, Xie T, Zafarmand MH, Zhao JH, Zhao W, Amin N, Arzumanyan Z, Astrup A, Bakker SJL, Baldassarre D, Beekman M, Bergman RN, Bertoni A, Bluher M, Bonnycastle LL, Bornstein SR, Bowden DW, Cai Q, Campbell A, Campbell H, Chang YC, de Geus EJC, Dehghan A, Du S, Eiriksdottir G, Farmaki AE, Franberg M, Fuchsberger C, Gao Y, Gjesing AP, Goel A, Han S, Hartman CA, Herder C, Hicks AA, Hsieh CH, Hsueh WA, Ichihara S, Igase M, Ikram MA, Johnson WC, Jorgensen ME, Joshi PK, Kalyani RR, Kandeel FR, Katsuya T, Khor CC, Kiess W, Kolcic I, Kuulasmaa T, Kuusisto J, Lall K, Lam K, Lawlor DA, Lee NR, Lemaitre RN, Li H, Lifelines Cohort Study, Lin SY, Lindstrom J, Linneberg A, Liu J, Lorenzo C, Matsubara T, Matsuda F, Mingrone G, Mooijaart S, Moon S, Nabika T, Nadkarni GN, Nadler JL, Nelis M, Neville MJ, Norris JM, Ohyagi Y, Peters A, Peyser PA, Polasek O, Qi Q, Raven D, Reilly DF, Reiner A, Rivideneira F, Roll K, Rudan I, Sabanayagam C, Sandow K, Sattar N, Schurmann A, Shi J, Stringham HM, Taylor KD, Teslovich TM, Thuesen B, Timmers P, Tremoli E, Tsai MY, Uitterlinden A, van Dam RM, van Heemst D, van Hylckama Vlieg A, van Vliet-Ostaptchouk JV, Vangipurapu J, Vestergaard H, Wang T, Willems van Dijk K, Zemunik T, Abecasis GR, Adair LS, Aguilar-Salinas CA, Alarcon-Riquelme ME, An P, Aviles-Santa L, Becker DM, Beilin LJ, Bergmann S, Bisgaard H, Black C, Boehnke M, Boerwinkle E, Bohm BO, Bonnelykke K, Boomsma DI, Bottinger EP, Buchanan TA, Canouil M, Caulfield MJ, Chambers JC, Chasman DI, Chen YI, Cheng CY, Collins FS, Correa A, Cucca F, de Silva HJ, Dedoussis G, Elmstahl S, Evans MK, Ferrannini E, Ferrucci L, Florez JC, Franks PW, Frayling TM, Froguel P, Gigante B, Goodarzi MO, Gordon-Larsen P, Grallert H, Grarup N, Grimsgaard S, Groop L, Gudnason V, Guo X, Hamsten A, Hansen T, Hayward C, Heckbert SR, Horta BL, Huang W, Ingelsson E, James PS, Jarvelin MR, Jonas JB, Jukema JW, Kaleebu P, Kaplan R, Kardia SLR, Kato N, Keinanen-Kiukaanniemi SM, Kim BJ, Kivimaki M, Koistinen HA, Kooner JS, Korner A, Kovacs P, Kuh D, Kumari M, Kutalik Z, Laakso M, Lakka TA, Launer LJ, Leander K, Li H, Lin X, Lind L, Lindgren C, Liu S, Loos RJF, Magnusson PKE, Mahajan A, Metspalu A, Mook-Kanamori DO, Mori TA, Munroe PB, Njolstad I, O’Connell JR, Oldehinkel AJ, Ong KK, Padmanabhan S, Palmer CNA, Palmer ND, Pedersen O, Pennell CE, Porteous DJ, Pramstaller PP, Province MA, Psaty BM, Qi L, Raffel LJ, Rauramaa R, Redline S, Ridker PM, Rosendaal FR, Saaristo TE, Sandhu M, Saramies J, Schneiderman N, Schwarz P, Scott LJ, Selvin E, Sever P, Shu XO, Slagboom PE, Small KS, Smith BH, Snieder H, Sofer T, Sorensen TIA, Spector TD, Stanton A, Steves CJ, Stumvoll M, Sun L, Tabara Y, Tai ES, Timpson NJ, Tonjes A, Tuomilehto J, Tusie T, Uusitupa M, van der Harst P, van Duijn C, Vitart V, Vollenweider P, Vrijkotte TGM, Wagenknecht LE, Walker M, Wang YX, Wareham NJ, Watanabe RM, Watkins H, Wei WB, Wickremasinghe AR, Willemsen G, Wilson JF, Wong TY, Wu JY, Xiang AH, Yanek LR, Yengo L, Yokota M, Zeggini E, Zheng W, Zonderman AB, Rotter JI, Gloyn AL, McCarthy MI, Dupuis J, Meigs JB, Scott RA, Prokopenko I, Leong A, Liu CT, Parker SCJ, Mohlke KL, Langenberg C, Wheeler E, Morris AP, Barroso I, Meta-Analysis of Glucose and Insulin-related Traits (MAGIC) Consortium. The trans-ancestral genomic architecture of glycemic traits. Nat Genet. 2021;53:840–860.

69. Graham SE, Clarke SL, Wu KH, Kanoni S, Zajac GJM, Ramdas S, Surakka I, Ntalla I, Vedantam S, Winkler TW, Locke AE, Marouli E, Hwang MY, Han S, Narita A, Choudhury A, Bentley AR, Ekoru K, Verma A, Trivedi B, Martin HC, Hunt KA, Hui Q, Klarin D, Zhu X, Thorleifsson G, Helgadottir A, Gudbjartsson DF, Holm H, Olafsson I, Akiyama M, Sakaue S, Terao C, Kanai M, Zhou W, Brumpton BM, Rasheed H, Ruotsalainen SE, Havulinna AS, Veturi Y, Feng Q, Rosenthal EA, Lingren T, Pacheco JA, Pendergrass SA, Haessler J, Giulianini F, Bradford Y, Miller JE, Campbell A, Lin K, Millwood IY, Hindy G, Rasheed A, Faul JD, Zhao W, Weir DR, Turman C, Huang H, Graff M, Mahajan A, Brown MR, Zhang W, Yu K, Schmidt EM, Pandit A, Gustafsson S, Yin X, Luan J, Zhao JH, Matsuda F, Jang HM, Yoon K, Medina-Gomez C, Pitsillides A, Hottenga JJ, Willemsen G, Wood AR, Ji Y, Gao Z, Haworth S, Mitchell RE, Chai JF, Aadahl M, Yao J, Manichaikul A, Warren HR, Ramirez J, Bork-Jensen J, Karhus LL, Goel A, Sabater-Lleal M, Noordam R, Sidore C, Fiorillo E, McDaid AF, Marques-Vidal P, Wielscher M, Trompet S, Sattar N, Mollehave LT, Thuesen BH, Munz M, Zeng L, Huang J, Yang B, Poveda A, Kurbasic A, Lamina C, Forer L, Scholz M, Galesloot TE, Bradfield JP, Daw EW, Zmuda JM, Mitchell JS, Fuchsberger C, Christensen H, Brody JA, Feitosa MF, Wojczynski MK, Preuss M, Mangino M, Christofidou P, Verweij N, Benjamins JW, Engmann J, Kember RL, Slieker RC, Lo KS, Zilhao NR, Le P, Kleber ME, Delgado GE, Huo S, Ikeda DD, Iha H, Yang J, Liu J, Leonard HL, Marten J, Schmidt B, Arendt M, Smyth LJ, Canadas-Garre M, Wang C, Nakatochi M, Wong A, Hutri-Kahonen N, Sim X, Xia R, Huerta-Chagoya A, Fernandez-Lopez JC, Lyssenko V, Ahmed M, Jackson AU, Yousri NA, Irvin MR, Oldmeadow C, Kim HN, Ryu S, Timmers P, Arbeeva L, Dorajoo R, Lange LA, Chai X, Prasad G, Lores-Motta L, Pauper M, Long J, Li X, Theusch E, Takeuchi F, Spracklen CN, Loukola A, Bollepalli S, Warner SC, Wang YX, Wei WB, Nutile T, Ruggiero D, Sung YJ, Hung YJ, Chen S, Liu F, Yang J, Kentistou KA, Gorski M, Brumat M, Meidtner K, Bielak LF, Smith JA, Hebbar P, Farmaki AE, Hofer E, Lin M, Xue C, Zhang J, Concas MP, Vaccargiu S, van der Most PJ, Pitkanen N, Cade BE, Lee J, van der Laan SW, Chitrala KN, Weiss S, Zimmermann ME, Lee JY, Choi HS, Nethander M, Freitag-Wolf S, Southam L, Rayner NW, Wang CA, Lin SY, Wang JS, Couture C, Lyytikainen LP, Nikus K, Cuellar-Partida G, Vestergaard H, Hildalgo B, Giannakopoulou O, Cai Q, Obura MO, van Setten J, Li X, Schwander K, Terzikhan N, Shin JH, Jackson RD, Reiner AP, Martin LW, Chen Z, Li L, Highland HM, Young KL, Kawaguchi T, Thiery J, Bis JC, Nadkarni GN, Launer LJ, Li H, Nalls MA, Raitakari OT, Ichihara S, Wild SH, Nelson CP, Campbell H, Jager S, Nabika T, Al-Mulla F, Niinikoski H, Braund PS, Kolcic I, Kovacs P, Giardoglou T, Katsuya T, Bhatti KF, de Kleijn D, de Borst GJ, Kim EK, Adams HHH, Ikram MA, Zhu X, Asselbergs FW, Kraaijeveld AO, Beulens JWJ, Shu XO, Rallidis LS, Pedersen O, Hansen T, Mitchell P, Hewitt AW, Kahonen M, Perusse L, Bouchard C, Tonjes A, Chen YI, Pennell CE, Mori TA, Lieb W, Franke A, Ohlsson C, Mellstrom D, Cho YS, Lee H, Yuan JM, Koh WP, Rhee SY, Woo JT, Heid IM, Stark KJ, Volzke H, Homuth G, Evans MK, Zonderman AB, Polasek O, Pasterkamp G, Hoefer IE, Redline S, Pahkala K, Oldehinkel AJ, Snieder H, Biino G, Schmidt R, Schmidt H, Chen YE, Bandinelli S, Dedoussis G, Thanaraj TA, Kardia SLR, Kato N, Schulze MB, Girotto G, Jung B, Boger CA, Joshi PK, Bennett DA, De Jager PL, Lu X, Mamakou V, Brown M, Caulfield MJ, Munroe PB, Guo X, Ciullo M, Jonas JB, Samani NJ, Kaprio J, Pajukanta P, Adair LS, Bechayda SA, de Silva HJ, Wickremasinghe AR, Krauss RM, Wu JY, Zheng W, den Hollander AI, Bharadwaj D, Correa A, Wilson JG, Lind L, Heng CK, Nelson AE, Golightly YM, Wilson JF, Penninx B, Kim HL, Attia J, Scott RJ, Rao DC, Arnett DK, Hunt SC, Walker M, Koistinen HA, Chandak GR, Yajnik CS, Mercader JM, Tusie-Luna T, Aguilar-Salinas CA, Villalpando CG, Orozco L, Fornage M, Tai ES, van Dam RM, Lehtimaki T, Chaturvedi N, Yokota M, Liu J, Reilly DF, McKnight AJ, Kee F, Jockel KH, McCarthy MI, Palmer CNA, Vitart V, Hayward C, Simonsick E, van Duijn CM, Lu F, Qu J, Hishigaki H, Lin X, Marz W, Parra EJ, Cruz M, Gudnason V, Tardif JC, Lettre G, t Hart LM, Elders PJM, Damrauer SM, Kumari M, Kivimaki M, van der Harst P, Spector TD, Loos RJF, Province MA, Psaty BM, Brandslund I, Pramstaller PP, Christensen K, Ripatti S, Widen E, Hakonarson H, Grant SFA, Kiemeney L, de Graaf J, Loeffler M, Kronenberg F, Gu D, Erdmann J, Schunkert H, Franks PW, Linneberg A, Jukema JW, Khera AV, Mannikko M, Jarvelin MR, Kutalik Z, Cucca F, Mook-Kanamori DO, van Dijk KW, Watkins H, Strachan DP, Grarup N, Sever P, Poulter N, Rotter JI, Dantoft TM, Karpe F, Neville MJ, Timpson NJ, Cheng CY, Wong TY, Khor CC, Sabanayagam C, Peters A, Gieger C, Hattersley AT, Pedersen NL, Magnusson PKE, Boomsma DI, de Geus EJC, Cupples LA, van Meurs JBJ, Ghanbari M, Gordon-Larsen P, Huang W, Kim YJ, Tabara Y, Wareham NJ, Langenberg C, Zeggini E, Kuusisto J, Laakso M, Ingelsson E, Abecasis G, Chambers JC, Kooner JS, de Vries PS, Morrison AC, North KE, Daviglus M, Kraft P, Martin NG, Whitfield JB, Abbas S, Saleheen D, Walters RG, Holmes MV, Black C, Smith BH, Justice AE, Baras A, Buring JE, Ridker PM, Chasman DI, Kooperberg C, Wei WQ, Jarvik GP, Namjou B, Hayes MG, Ritchie MD, Jousilahti P, Salomaa V, Hveem K, Asvold BO, Kubo M, Kamatani Y, Okada Y, Murakami Y, Thorsteinsdottir U, Stefansson K, Ho YL, Lynch JA, Rader DJ, Tsao PS, Chang KM, Cho K, O’Donnell CJ, Gaziano JM, Wilson P, Rotimi CN, Hazelhurst S, Ramsay M, Trembath RC, van Heel DA, Tamiya G, Yamamoto M, Kim BJ, Mohlke KL, Frayling TM, Hirschhorn JN, Kathiresan S, V. A. Million Veteran Program, Global Lipids Genetics Consortium, Boehnke M, Natarajan P, Peloso GM, Brown CD, Morris AP, Assimes TL, Deloukas P, Sun YV and Willer CJ. The power of genetic diversity in genome-wide association studies of lipids. Nature. 2021;600:675–679.

70. Mehrabian M, Schulthess FT, Nebohacova M, Castellani LW, Zhou Z, Hartiala J, Oberholzer J, Lusis AJ, Maedler K and Allayee H. Identification of ALOX5 as a gene regulating adiposity and pancreatic function. Diabetologia. 2008;51:978–88.

71. Woodward NC, Crow AL, Zhang Y, Epstein S, Hartiala J, Johnson R, Kocalis H, Saffari A, Sankaranarayanan I, Akbari O, Ramanathan G, Araujo JA, Finch CE, Bouret SG, Sioutas C, Morgan TE and Allayee H. Exposure to nanoscale particulate matter from gestation to adulthood impairs metabolic homeostasis in mice. Sci Rep. 2019;9:1816.

72. Matthews DR, Hosker JP, Rudenski AS, Naylor BA, Treacher DF and Turner RC. Homeostasis model assessment: insulin resistance and beta-cell function from fasting plasma glucose and insulin concentrations in man. Diabetologia. 1985;28:412–9.

73. Mehrabian M, Allayee H, Wong J, Shi W, Wang XP, Shaposhnik Z, Funk CD and Lusis AJ. Identification of 5-lipoxygenase as a major gene contributing to atherosclerosis susceptibility in mice. Circ Res. 2002;91:120–6.

